# A data driven approach to assess relationships between sleep, cognition and dementia: Findings from the Sleep and Dementia Consortium

**DOI:** 10.64898/2025.12.17.25342519

**Authors:** Stephanie Yiallourou, Crystal Wiedner, Qiong Yang, Andree-Ann Baril, Jeffrey R. Misialek, Christopher E. Kline, Stephanie Harrison, Rebecca Bernal, Alycia Bisson, Dibya Himali, Tobyn Chiu, Marina Cavuoto, Sonia Ancoli-Israel, Qian Xiao, Eleni Okeanis Vaou, Antoine Weihs, Yue Leng, Rebecca F. Gottesman, Alexa Beiser, Oscar Lopez, Pamela L. Lutsey, Shaun M. Purcell, Susan Redline, Sudha Seshadri, Katie L. Stone, Kristine Yaffe, Matthew P. Pase, Jayandra J. Himali

## Abstract

**Background and Objectives:** Sleep has been associated with cognition and risk of dementia. However, sleep is a highly complex and multi-dimensional state, and there is uncertainty about which aspects of sleep are most relevant to cognitive performance and dementia risk. We applied a data-driven approach to identify clusters of sleep variables that reflect meaningful sleep composites and examined their association with cognitive performance and dementia risk.

**Methods:** Data from the Sleep and Dementia Consortium, consisting of 5 US population-based cohorts were utilized. Participants had methodologically consistent, home-based polysomnography, self-report habitual sleep, neuropsychological assessments, and dementia risk surveillance. The pooled cognitive analysis included 5,958 participants aged ≥45 years, and the incident dementia analysis included 5,471 participants aged ≥60 years. A cluster around latent variables analysis was used to derive 9 latent sleep composites from 44 sleep metrics. Global cognitive composite z-scores were derived from principal component analysis. Linear regression models were used to assess associations between sleep composites and cognitive performance. Cox proportional hazard models assessed associations between sleep composites and incident dementia.

**Results:** Mean (SD) age was 70 ± 11 and 74 ± 12 years for the cognitive and dementia analysis, respectively. There were 1,134 incident dementia cases (median follow-up time of 5-19 years). 9 sleep composites were identified, together explaining 49% of the total variance in the original 44 sleep metrics: Sleep quantity and efficiency, sleep fragmentation, light NREM predominance, N3 predominance, spindle number and duration, REM sleep bouts, respiratory disturbances, slow oscillation-spindle coupling and spindle amplitude. Of these, composites reflecting greater sleep quantity and efficiency (i.e., longer and more consolidated sleep; pooled β per one-unit change in composite, 0.03; 95% CI: 0.004 - 0.06; p=0.033) and stronger slow oscillation-spindle coupling (pooled β, 0.04; 95% CI: 0.003 - 0.07; p=0.039) were associated with better global cognition. However, no significant associations were identified between the 9 sleep composites and dementia risk.

**Discussion:** Our data-driven approach identified longer, more consolidated sleep and stronger slow oscillation-spindle coupling as the composites of sleep most strongly related to cognitive performance. These composites may be useful in guiding further investigations of sleep-brain health relationships.

## INTRODUCTION

Sleep is important for cognitive function^1^ and is thought to play a vital role in the clearance of neurotoxic proteins that accumulate in neurodegenerative disorders^2^. Animal studies demonstrate that clearance of amyloid-β, a key protein that accumulates in Alzheimer’s disease, is highest during sleep and coupled to slow wave sleep (Stage 3 sleep, N3)^2^. Accordingly, sleep may be an important target for dementia risk reduction.

Though sleep dysfunction has been linked to amyloid accumulation^3^, studies investigating sleep and dementia associations have yielded mixed results. Several studies have shown that short^4–6^ or long sleep duration^7^ is associated with dementia risk, while other have shown no association^8, 9^. In the largest study to date, utilizing curated data from 5,946 adults across five U.S. cohorts that form the Sleep and Dementia Consortium (SDC), we identified that obstructive sleep apnea (OSA) (across the severity spectrum) was associated with poorer global cognitive function, while higher sleep maintenance efficiency was associated with better cognitive performance^10^. However, in these cohorts, there was little evidence to suggest that these same sleep metrics were related to dementia risk^9^. Notably, the amount of N3 or sleep duration was not robustly associated with cognition or dementia risk^9^.

The lack of consistent sleep and dementia associations may arise due to the complexity of conceptualizing and measuring sleep. Specifically, individual metrics may not fully capture the neurophysiological intricacies of sleep that contribute to cognition and dementia risk. The majority of previous research has been restricted to exploring sleep by self-report measures (e.g., sleep duration)^11^ or by objective summary measures of sleep architecture and sleep duration. (e.g., organization of sleep stages or objective sleep duration)^12^. However, sleep is a highly dynamic, multi-dimensional, and complex physiological state, with a vast array of sleep metrics available to characterize differing aspects of its macro- and micro-architecture, timing, and continuity. Furthermore, many of these metrics may have interrelated properties. Exactly which sleep metrics to target in dementia research remains unresolved, with most studies considering many measures in isolation, which leads to issues of multiple comparisons and potential errors in inference.

To overcome these limitations, we applied a data-driven approach, performing clustering around latent variables analyses (CLV).^13^ This allowed us to comprehensively assess a full spectrum of sleep metrics (e.g., sleep macro- and micro-architecture, respiratory disturbance, and self-reported sleep quality) and reduce these sleep metrics into a smaller number of biologically meaningful latent sleep composites. Using data from the SDC, we aimed to derive clusters of sleep variables (latent sleep composites) and identify which of these were most strongly associated with cognitive performance and dementia risk.

## METHODS

### Standard Protocol Approvals, Registrations, and Patient Consents

All participants in the SDC provided written informed consent before study commencement. Each cohort obtained institutional review board (IRB) approval at its respective institution, and this study was approved by the IRB at the University of Texas Health Science Center at San Antonio. This report follows the Strengthening the Reporting of Observational Studies in Epidemiology (STROBE) reporting guideline for cohort studies.

### Study Design

The SDC comprises 5 prospective community-based cohorts that have performed methodologically consistent, overnight, home-based polysomnography (PSG) processed and scored at a single sleep reading center. All cohorts also underwent neurocognitive assessments and dementia case ascertainment. Cohorts include the Atherosclerosis Risk in Communities (ARIC) study https://www5.cscc.unc.edu/aric9/, Cardiovascular Health Study (CHS) https://chs-nhlbi.org/, Framingham Heart Study (FHS) https://www.framinghamheartstudy.org, Osteoporotic Fractures in Men Study (MrOS) https://mrosonline.ucsf.edu, and Study of Osteoporotic Fractures (SOF) https://sofonline.ucsf.edu. For this analysis, we investigated associations between baseline sleep composites and 1) cognitive function measured within the subsequent 5 years and 2) dementia risk with up to a median of 19.2 years of follow-up. The timing of assessments is shown in the Supplement, eTable 1.

### Participants

Enrolment and cohort design methods for the SDC have been published previously^10^. The cognitive analysis was limited to participants aged at least 45 years who were free of dementia and stroke and who had baseline PSG and cognitive testing within 5 years after PSG. The dementia analysis was limited to participants aged ≥60 years (to limit cases to late onset dementia) at the time of PSG, free of dementia and other neurological disorders (other neurological conditions, including stroke, multiple sclerosis, significant head trauma, subdural hematoma, brain tumor, etc.) at the baseline PSG, and who had information on dementia status after PSG. To reduce the influence of potentially inaccurate data, participants with less than 180 minutes of total sleep time or less than 1 minute of REM sleep were excluded. For CHS, analyses were limited to those with available DNA who consented to genetic studies. Sample selection across cohorts is shown in eTable 2.

### Sleep Assessment

#### Polysomnography (PSG)

Participants in the ARIC, CHS, and FHS cohorts completed in-home PSG as part of the Sleep Heart Health Study^14^. For SOF and MrOS, sleep protocols were modelled on the Sleep Heart Health Study, enabling effective harmonization and cross-study comparison. All cohorts used a standardized protocol to complete overnight, home-based type II PSGs between 1995 and 1998 for ARIC, CHS, and FHS (Compumedics P Series System, Abbotsford) and 2000 to 2004 for MrOS (Compumedics Safiro Unit) and SOF (Compumedics Siesta Unit). Electroencephalogram (C3A2 and C4A1), electrooculogram, electromyogram, thoracic and abdominal displacement (inductive plethysmography bands), airflow (nasal-oral thermocouples), finger pulse oximeter, a single bipolar electrocardiogram, body position by a mercury gauge sensor, and ambient light level were all recorded. SOF and MrOS added nasal cannula pressure and bilateral piezoelectric sensors to detect leg movements.

The same central Sleep Reading Center oversaw training, conduct, and analysis of all sleep studies. Methods, including scoring guidelines and reliability, have been previously published^14–16^. Sleep was scored in 30-second epochs by trained PSG technicians blinded to other data according to established guidelines (Rechtschaffen and Kales^17^ and American Sleep Disorders Association arousal criteria^18^).

Measures of sleep macroarchitecture and respiratory disturbance were derived from manual scoring. Measures of sleep stage transitions, EEG slow oscillation and spindle morphology metrics, and sleep bout durations were obtained via the automated LUNA software. In addition to PSG, we also included self-reported habitual sleep duration (hours) and daytime sleepiness assessed via the Epworth Sleepiness Scale^19^. In total, 44 sleep metrics were derived and are listed in Table 1. The supplementary methods provides a detailed description of how these metrics were assessed.

**Table 1:**
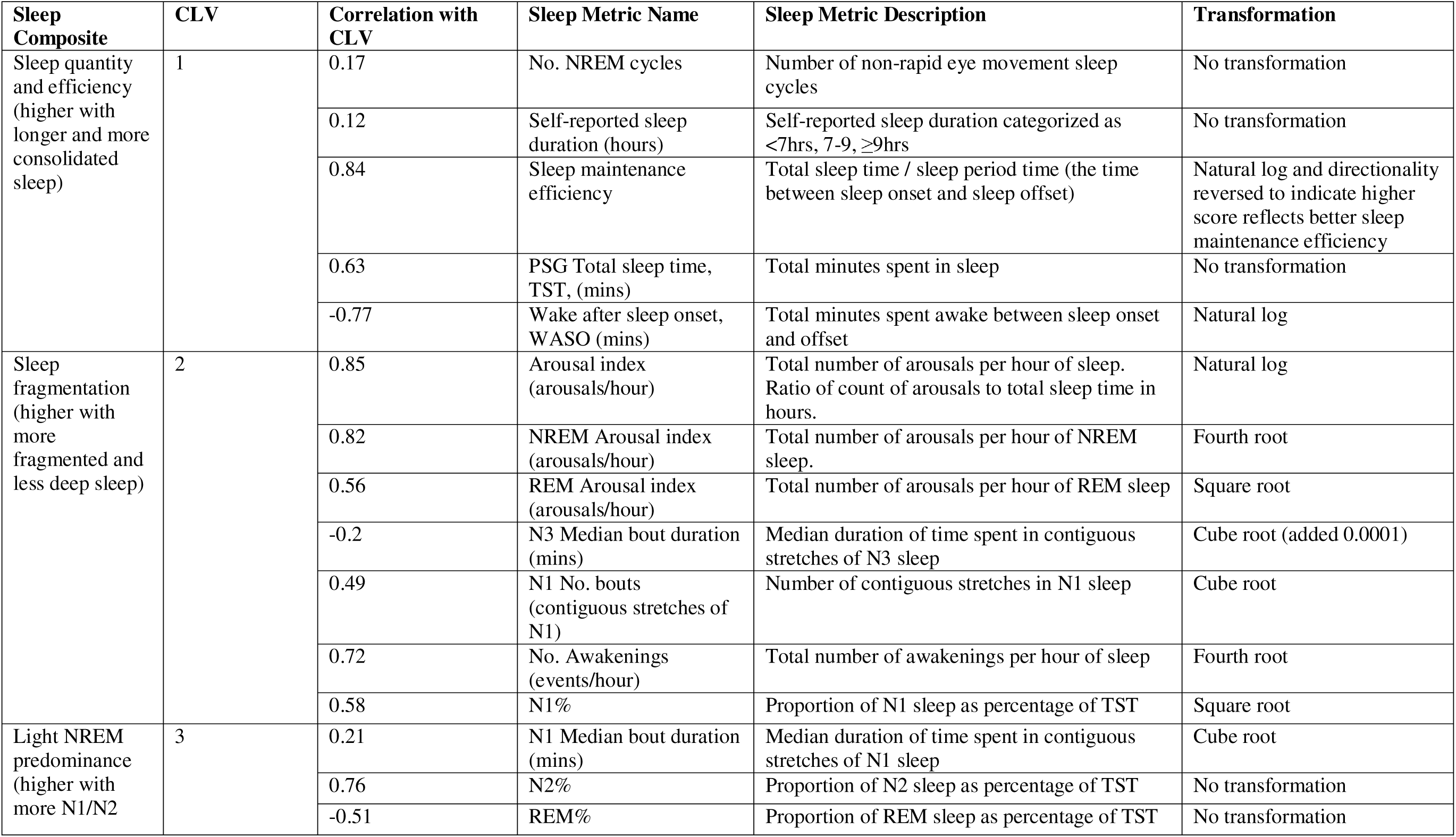
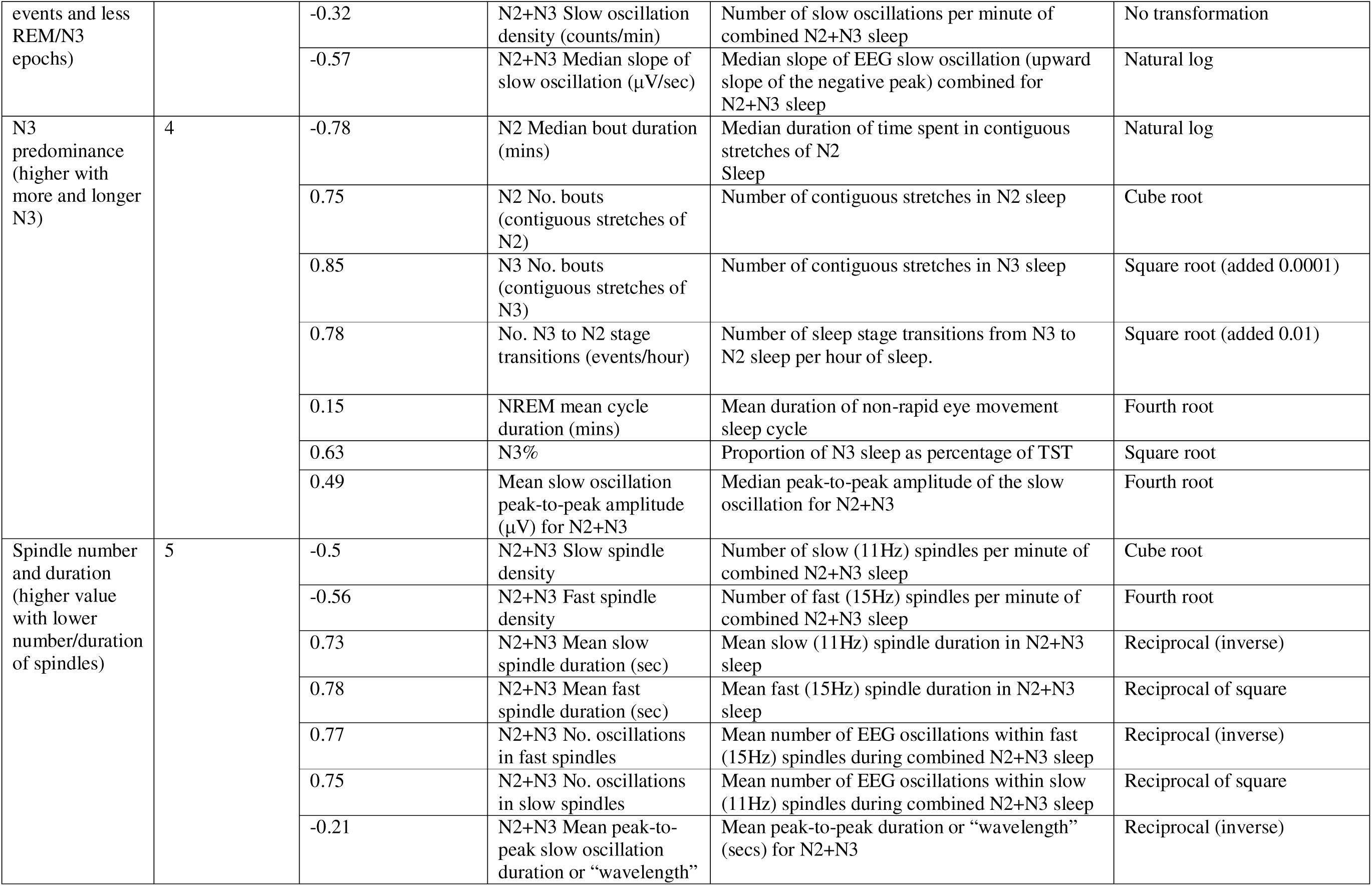
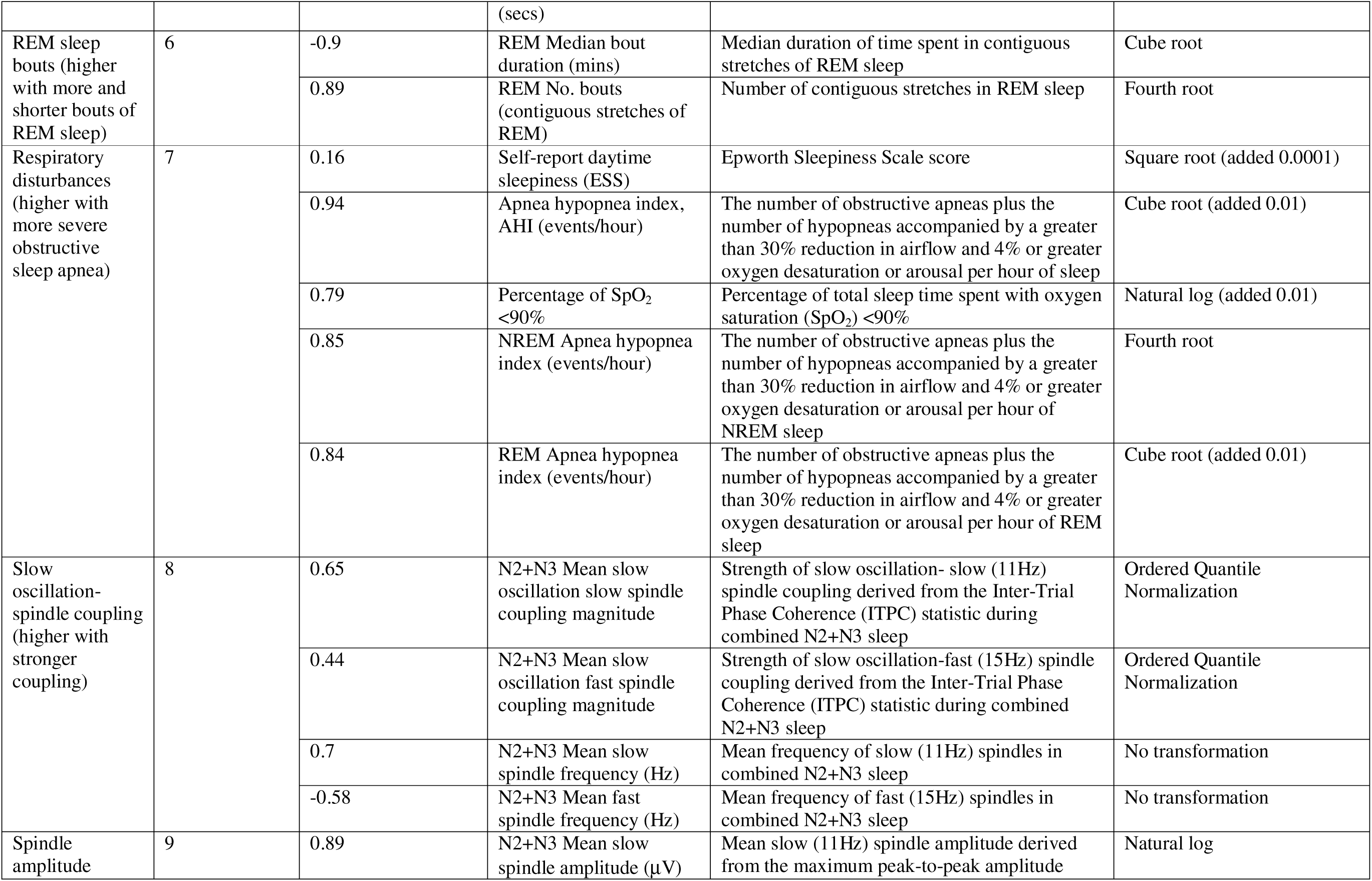
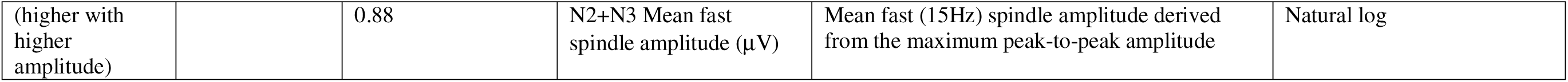
9 Sleep composites identified from clustering around latent variables (CLV) analyses.

### Cognitive Assessment

Cognition was assessed within 5 years of the sleep study across all cohorts (average time between PSG and cognitive assessment: 0.3 to 4.6 years) The methods used to assess cognition varied between cohorts and have been published previously^10^. Performance on any given cognitive test partly depends on a general cognitive ability, and studies have shown that a single cognitive factor can explain more than 40% variance across test batteries^20^. Thus, to allow for a consistent measure of general cognition, even when tests differed across studies, a general cognitive score was calculated from at least three different cognitive tests for each participant within each cohort^21^. To derive this score, principal component analysis was applied to the cognitive test scores, forcing a single-factor solution. Individual tasks to create the global cognitive composite for each cohort and their factor loadings have been published previously^10^ and are summarized in eTable 3.

In a secondary analysis, cognitive tests were grouped into broader cognitive domains, including executive function, attention, processing speed, verbal learning and memory, language, and visuospatial ability. Test scores were converted to z-scores based on the mean and standard deviation within each cohort. Any tests where higher scores indicated poorer performance were transformed (by negative 1) so that higher scores consistently indicated better cognitive performance.

### Dementia Case Ascertainment

Dementia case ascertainment has been described in detail^9, 10^. In brief, ARIC, CHS, FHS, and SOF adjudicated dementia diagnosis using different combinations of neurocognitive assessments, informant interviews, and hospitalization records, based broadly on the Diagnostic and Statistical Manual of Mental Disorders 4^th^ or 5^th^ edition (DSM-IV/V) criteria. MrOS investigators adjudicated clinically significant cognitive impairment by a report of physician-diagnosed dementia, use of dementia medication, or a change in modified Mini-Mental State Examination scores ≥ 1.5 standard deviations worse than the mean change from baseline to any follow-up visit. FHS conducted continuous surveillance of dementia, with potential cases adjudicated by a committee according to DSM-IV/V criteria (or equivalent). CHS also had a committee adjudicate dementia cases according to the DSM-IV, as described previously^9^. For ARIC, cases were identified by hospitalization and death ICD-9/10 codes from study baseline until the end of follow-up^22^. Beginning in 2011, this approach was complemented by intensive in-clinic cognitive testing and phone surveillance. Both MrOS and SOF assessed dementia at discrete follow-up time points (e.g., several years), up to approximately 10 and 5 years following sleep assessment, respectively.

### Covariates

The following covariates were assessed at the time of PSG and included in statistical models: age (years), sex (male vs. female), body mass index (kg/m^2^), anti-depressant use (yes vs. no), sedative use (yes vs. no). The models involving cognitive outcomes included additional adjustments for age^2^ (years), education (<high school [<12 years], high school [12 years], and >high school [>12 years]), and the time interval between the PSG assessment and cognitive testing (years).

### Statistical Analysis

Statistical analysis was performed using SAS version 9.4 (SAS Institute Inc., Cary, NC, USA) and R version 4.3.0 (R Foundation for Statistical Computing, Vienna, Austria). Demographic characteristics were examined by study cohort.

#### Clustering around latent variables (CLV) analyses

A detailed explanation of the methods used to reduce sleep metrics into sleep composites is provided in the supplement. Briefly, the 44 sleep metrics were selected to provide a broad representation of sleep indices, based on their known relevance to cognition and dementia, and their consistency across all cohorts. CLV methods^13^ were used on the 44 sleep metrics to identify meaningful clusters, where correlated sleep metrics were grouped together and a latent (synthetic) variable was formed based on sleep metrics within the same cluster. The age-, sex-, and study center-adjusted correlations (Supplementary eFigure 1) were used in the cluster analyses. CLV employed an agglomerative technique, starting with as many clusters as the number of variables (each variable was a cluster by itself). Then in the next iteration, the number of clusters was reduced by one by aggregating two existing clusters that were chosen based on the minimum reduction of the sum of maximum eigenvalues of each cluster over all clusters before and after merging the two clusters. The iterations continued until reaching a single cluster of all variables. We started with the number of clusters (4) after aggregation that corresponds to the first large jump in the elbow plot of merging criterion as final clusters (Supplementary eFigure 2). Then, we examined the results when choosing an adjacent number of clusters as cutoffs by evaluating the variance of original variables explained by all the clusters, individual cluster variances, and members of each cluster. We then chose the 9 cluster configurations, such that 49% variance was explained by the latent variables. Once complete, we used expert knowledge to name each cluster (sleep composites).

**Figure 1:**
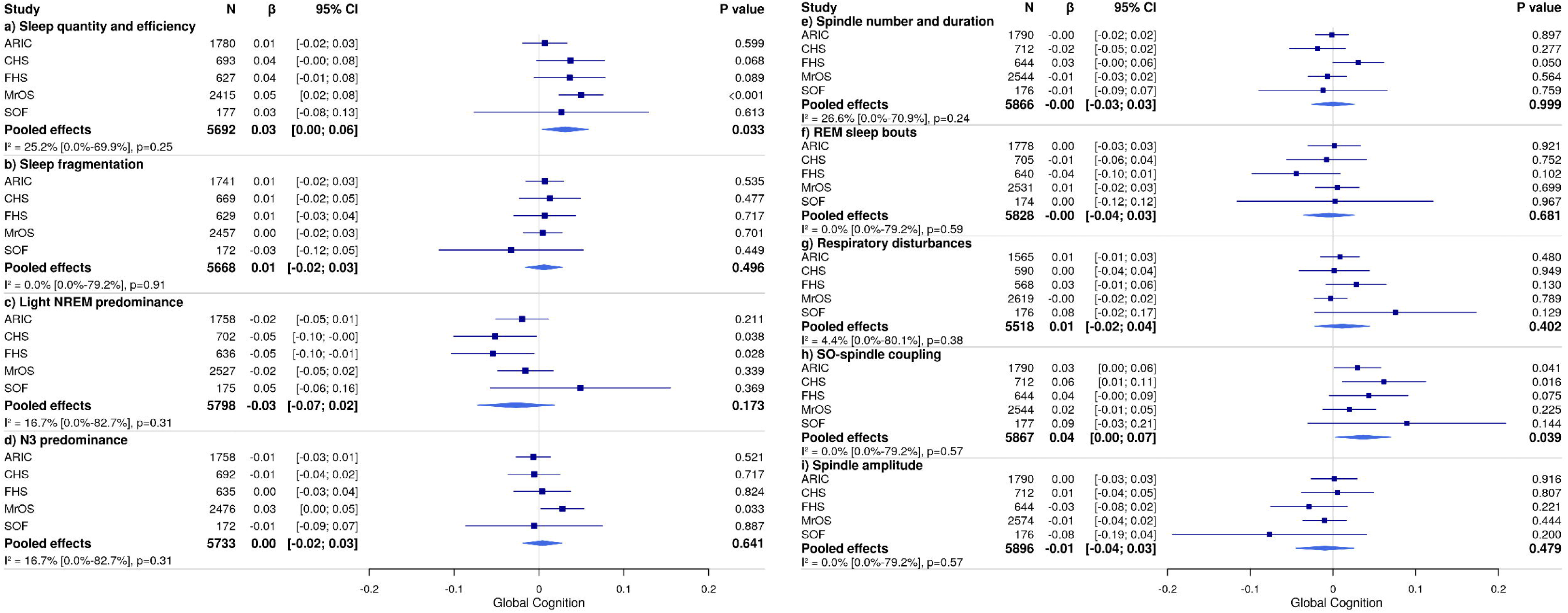
Forest plots of the associations between sleep composites and global cognition. Models were adjusted for age, age^2^, sex, body mass index, anti-depressant use, sedative use, education, and the time interval between the PSG assessment and cognitive testing. The directionality for each sleep composite was interpreted as: Sleep quantity and efficiency = higher with longer and more consolidated sleep; Sleep fragmentation = higher with more fragmented sleep; Light NREM predominance = higher with more N1/N2 events and less REM/N3 events; N3 predominance = higher with more and longer N3; Spindle number and duration = higher value with lower number/ duration of spindles; REM sleep bouts = higher with more and shorter bouts of REM sleep; Respiratory disturbances = higher with more OSA); Spindle and slow wave coupling = higher with stronger coupling; Spindle amplitude = higher with higher amplitude. Abbreviations: NREM, non-rapid eye movement sleep; N1, stage 1 non-rapid eye movement sleep; N2, stage 2 non-rapid eye movement sleep; N3, non-rapid eye movement sleep; REM, rapid eye movement sleep; OSA, obstructive sleep Apnea; ARIC, Atherosclerosis Risk in Communities study; CHS, Cardiovascular Health Study; FHS, Framingham Heart Study; MrOS, Osteoporotic Fractures in Men Study; SOF, Study of Osteoporotic Fractures.

**Figure 2:**
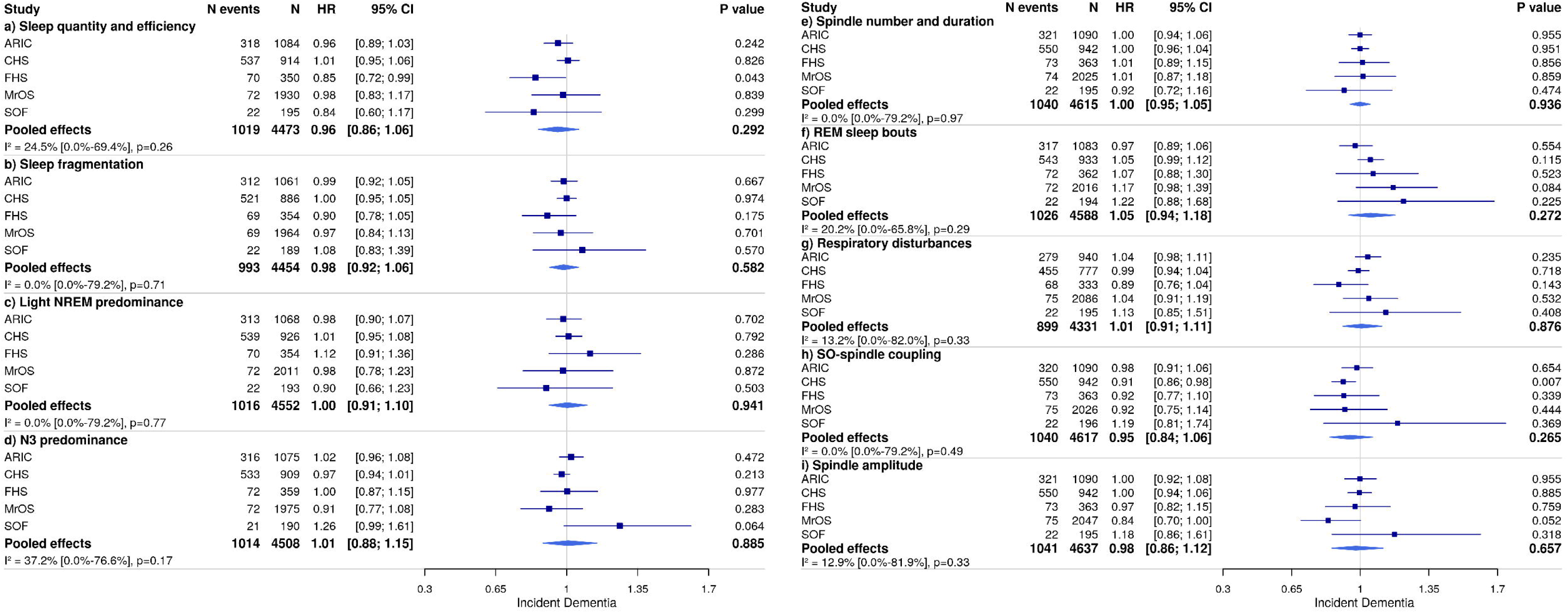
Forest plots of the associations between sleep composites and dementia risk. Models were adjusted for age, sex, body mass index, anti-depressant use, sedative use, and APOE *e4* status. The directionality for each sleep composite was interpreted as: Sleep quantity and efficiency = higher with longer and more consolidated sleep; Sleep fragmentation = higher with more fragmented sleep; Light NREM predominance = higher with more N1/N2 events and less REM/N3 events; N3 predominance = higher with more and longer N3; Spindle number and duration = higher value with lower number/ duration of spindles; REM sleep bouts = higher with more and shorter bouts of REM sleep; Respiratory disturbances = higher with more OSA); Spindle and slow wave coupling = higher with stronger coupling; Spindle amplitude = higher with higher amplitude. Abbreviations: NREM, non-rapid eye movement sleep; N1, stage 1 non-rapid eye movement sleep; N2, stage 2 non-rapid eye movement sleep; N3, non-rapid eye movement sleep; REM, rapid eye movement sleep; OSA, obstructive sleep Apnea; ARIC, Atherosclerosis Risk in Communities study; CHS, Cardiovascular Health Study; FHS, Framingham Heart Study; MrOS, Osteoporotic Fractures in Men Study; SOF, Study of Osteoporotic Fractures.

#### Cognitive and dementia analyses

Cohort-specific analyses were conducted for both cognitive and dementia outcomes. To determine the association between the 9 sleep composites and cognitive performance, separate linear regression models were applied, adjusting for the covariates listed above. Cox proportional hazard regression models were used to examine the association between the 9 sleep composites and incident all-cause dementia by estimating hazard ratios (HRs) with 95% confidence intervals (CIs). For dementia risk, we ran two models. To maximize sample size, we first analyzed data with a primary model (Model 1) that adjusted for age and sex. We then examined associations with a fully adjusted model (Model 2) which included all the covariates listed in the covariate section with the addition of APOE *e4* status (non-*e4* carrier vs. at least one copy of the *e4* allele). Missing data were excluded from analyses. Follow-up date ranges for each cohort are provided in the Supplementary Methods eTable1, and follow-up median durations are provided in Table 3. For each cohort, dementia follow-up times are displayed in eTable1. Non-events were censored at death or until the last date they were known to be dementia-free, or until administrative censoring. The proportional hazard assumption was examined by including an interaction term between the sleep exposure and the log of follow-up time. The assumption was confirmed graphically and statistically (p-value>0.05) in all cohorts.

#### Meta-analyses

Study-level estimates were pooled centrally in random effects meta-analyses. The Sidik-Jonkman estimator method was used to calculate the heterogeneity variance τ^2^, and the Hartung and Knapp method was used to calculate the 95% CIs around the pooled effect. The Higgins I^2^ test was implemented to test for heterogeneity in effect sizes.^23^ Statistical tests were all two-sided. All results were considered significant if p<0.05.

#### Effect Modification

In cohorts where data were available, we further explored effect modification by sex (male vs female) (ARIC, CHS and FHS cohorts) and APOE *e4* allele carrier status (carrier versus non-carrier) (ARIC, CHS, FHS, MrOS and SOF cohorts) for both cognitive and dementia risk outcomes in model 1. The interaction results were pooled and meta-analyses were performed.

### Data Availability

Deidentified participant data are available to qualified researchers whose proposed use has been approved. For the ARIC study, data are available upon reasonable request and subject to approval by the study investigators. Sleep data can be requested from the National Sleep Research Resource (NSRR; https://sleepdata.org/), and cognitive data can be requested from each participating cohort or from repositories such as BioLINCC (https://biolincc.nhlbi.nih.gov/home/). Data may be used for any purpose pending site approval. Access is provided after proposal approval or with a signed data access agreement.

## RESULTS

### Participant Characteristics

Sample characteristics for each cohort are presented in Table 2. In the cognitive outcome sample, there were 5,958 participants in total. While ARIC, CHS, and FHS comprised just over 50% women, the pooled sample comprised only 31.5% women due to the large all-male MrOS cohort. The mean age of participants at the time of PSG varied across the cohorts, with FHS being the youngest at 59 years, and SOF being the oldest, almost 82 years. Over 90% of the participants identified as White.

**Table 2.** Cohort characteristics at the time of the sleep study – cognitive sample.

|  | ARIC<br>(n=1791) | CHS<br>(n=712) | FHS<br>(n=644) | MrOS (n=2619) | SOF<br>(n=192) |
| --- | --- | --- | --- | --- | --- |
| Age, years | 62.4 (5.7) | 76.8 (3.8) | 59 (8.4) | 76.2 (5.5) | 81.9 (2.9) |
| 45-54 | 140 (7.8) | 0 (0.0) | 230 (35.7) | 0 (0.0) | 0 (0.0) |
| 55-64 | 983 (54.9) | 0 (0.0) | 230 (35.7) | 0 (0.0) | 0 (0.0) |
| 65-74 | 665 (37.1) | 200 (28.1) | 160 (24.8) | 1153 (44.0) | 0 (0.0) |
| 75-84 | 3 (0.2) | 487 (68.4) | 24 (3.7) | 1253 (47.8) | 28 (14.6) |
| 85-94 | 0 (0.0) | 25 (3.5) | 0 (0.0) | 211 (8.1) | 161 (83.9) |
| 95+ | 0 (0.0) | 0 (0.0) | 0 (0.0) | 2 (0.1) | 3 (1.6) |
| Female, n (%) | 927 (51.8) | 411 (57.7) | 345 (53.6) | 0 (0.0) | 192 (100.0) |
| Race |  |  |  |  |  |
| White, n (%) | 1778 (99.3) | 609 (85.5) | 520 (80.8) | 2382 (91.0) | 176 (91.7) |
| Black, n (%) | 7 (0.4) | 102 (14.3) | 54 (8.4) | 81 (3.1) | 15 (7.8) |
| Other, n (%) | 6 (0.3) | 1 (0.1) | 70 (10.9) | 156 (5.9) | 1 (0.5) |
| Education |  |  |  |  |  |
| Less than high school [<12 years], n (%) | 185 (10.3) | 128 (18.0) | 48 (7.5) | 114 (4.4) | 31 (17.5) |
| High school [12], n (%) | 641 (35.8) | 396 (55.6) | 173 (26.9) | 420 (16.0) | 88 (49.7) |
| More than high school [>12], n (%) | 965 (53.9) | 188 (26.4) | 423 (65.7) | 2085 (79.6) | 58 (32.8) |
| Time interval between PSG and cognitive testing, years | 0.3 (0.8) | 2.1 (0.6) | 3.7 (1.4) | 0 (0.0) | 4.6 (0.7) |
| Systolic blood pressure, mmHg | 121 (17.2) | 130 (18.7) | 125 (17.3) | 127 (16.1) | 136 (16.7) |
| Treatment for hypertension, n (%) | 606 (33.8) | 388 (54.5) | 159 (24.7) | 1735 (66.3) | 137 (71.4) |
| Stage I Hypertension, n (%) | 632 (35.3) | 400 (56.2) | 244 (38.0) | 1876 (71.6) | 156 (81.7) |
| Prevalent diabetes, n (%) | 215 (12) | 83 (11.69) | 31 (5.0) | 342 (13.1) | 18 (9.4) |
| Prevalent CVD, n (%) | 282 (15.8) | 72 (10.1) | 42 (6.5) | 1032 (39.5) | 36 (20.3) |
| Smoking status, n (%) | 176 (9.8) | 41 (5.8) | 85 (13.2) | 51 (1.9) | 3 (1.6) |
| Body mass index, Median [Q1, Q3] | 28.3 [25.3, 31.7] | 27.4 [24.9, 29.8] | 27.6 [25.0, 31.2] | 26.7 [24.6, 29.3] | 27.6 [25.2, 31.4] |
| Sleeping pill use, n (%) | 434 (24.2) | 159 (22.7) | 117 (18.5) | 362 (13.8) | 37 (19.3) |
| Antidepressant use, n (%) | 128 (7.2) | 35 (4.9) | 38 (5.9) | 192 (7.3) | 15 (7.8) |
| Sedative use, n (%) | 86 (4.8) | 47 (6.6) | 24 (3.7) | 183 (7.0) | 28 (14.6) |
| APOE ε4 allele, n (%) | 483 (27.0) | 157 (22.1) | 137 (21.7) | 497 (23.6) | 7 (7.7) |

Abbreviations: ARIC, Atherosclerosis Risk in Communities study; CHS, Cardiovascular Health Study; FHS, Framingham Heart Study; MrOS, Osteoporotic Fractures in Men Study; SOF, Study of Osteoporotic Fractures.

Study characteristics for the dementia analysis sample are presented in eTable 4. Sample characteristics were similar to the cognitive sample except that, by design, the dementia sample was older, with a mean age of 66 to 83 years across the cohorts. In total, there were 1,134 (20.7%) incident dementia cases across cohorts. The median follow-up time ranged between 4.5 to 19.2 years across cohorts (Table 3).

**Table 3.**
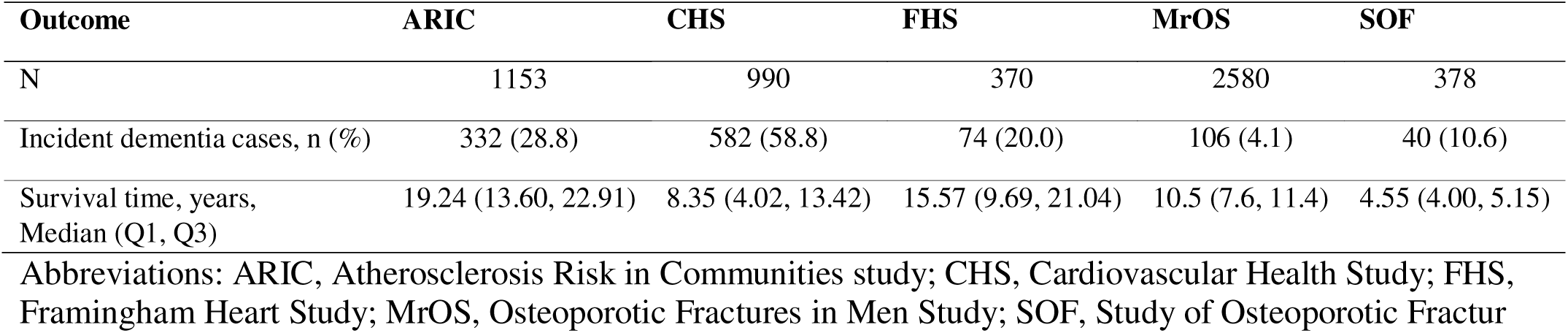
Sample for incident dementia analysis.

| Outcome | ARIC | CHS | FHS | MrOS | SOF |
| --- | --- | --- | --- | --- | --- |
| N | 1153 | 990 | 370 | 2580 | 378 |
| Incident dementia cases, n (%) | 332 (28.8) | 582 (58.8) | 74 (20.0) | 106 (4.1) | 40 (10.6) |
| Survival time, years,<br>Median (Q1, Q3) | 19.24 (13.60, 22.91) | 8.35 (4.02, 13.42) | 15.57 (9.69, 21.04) | 10.5 (7.6, 11.4) | 4.55 (4.00, 5.15) |

Abbreviations: ARIC, Atherosclerosis Risk in Communities study; CHS, Cardiovascular Health Study; FHS, Framingham Heart Study; MrOS, Osteoporotic Fractures in Men Study; SOF, Study of Osteoporotic Fractur

### Cluster of Variables around Latent Components (CLV): Sleep Composites

Table 1 presents sleep composites identified by the CLV procedures. Nine clusters explaining 49% of the variance in the original individual variables were selected, each containing variables meaningfully classified to a conceptually interpretable sleep composite. Within each cluster, the variance explained by the latent variable (i.e., the first principal component of variables in the cluster) ranged from 31% to 91% (median=54.5%).

### Associations between Sleep Composites and Global Cognition

Pooled estimates between the 9 sleep composites and global cognition are presented in Figure 1. Higher levels of sleep quantity and efficiency (pooled β, 0.03; 95% CI, 0.004 to 0.06; p-value = 0.033) and stronger slow oscillation-spindle coupling (pooled β, 0.04; 95% CI, 0.003 to 0.07; p-value = 0.039) were associated with better global cognition. No other pooled associations reached statistical significance. For the N3 predominance composite, more and longer N3 sleep was associated with better cognition in the MrOS cohort only (β, 0.03; 95% CI, 0.002 to 0.05; p-value = 0.033), but no other associations were found.

To explore which sleep metrics specifically drove the associations between sleep quantity and efficiency and slow oscillation-spindle coupling composites and global cognition, we analyzed the composite components separately. Results are presented in eFigures 3 and 4. Within the sleep quantity and efficiency composite, sleep maintenance efficiency and wake after sleep onset showed the strongest (albeit nonsignificant) trends with global cognition. This suggests that the latent construct of sleep quantity and efficiency captures shared variance across metrics, with some components contributing more strongly than others, yielding an association not apparent when metrics were examined in isolation. Within the slow oscillation-spindle coupling composite, the slow oscillation-fast spindle coupling magnitude (pooled β, 0.03; 95% CI, 0.002 to 0.05; p-value = 0.041) and the slow spindle frequency (pooled β, 0.16; 95% CI, 0.01 to 0.31; p-value = 0.039) were associated with higher global cognitive scores.

### Associations between Sleep Composites and Cognitive Domains

The associations between the sleep and individual cognitive composites are presented in eTable 5. The pooled estimates did not reveal any significant associations for any of the cognitive domains explored.

### Associations between Sleep Composites and Incident All-cause Dementia

Results did not differ meaningfully between model 1 and model 2, thus we present findings for the fully adjusted model 2 and have provided model 1 results in the supplementary material (eFigure 5). Figure 2 shows the pooled hazard ratio estimates for each sleep composite. Pooled estimates revealed no significant associations for any of the 9 sleep composites. However, higher scores on the sleep quantity and efficiency composite were associated with reduced dementia risk in the FHS cohort only (hazard ratio, 0.85; 95% CI 0.72 to 0.99; p=0.043). Further, stronger slow oscillation-spindle coupling was associated with reduced dementia risk in the CHS cohort (hazard ratio, 0.91; 95% CI 0.86 to 0.98; p=0.007).

### Examination of Interactions

The pooled effects were not suggestive of any consistent sex- or APOE- by sleep composite interactions (eTables 6-9).

## DISCUSSION

We harmonized 44 sleep metrics across 5 U.S.-based cohort studies and reduced these into 9 meaningful sleep composites that capture unique sleep processes. Our analysis revealed that better sleep quantity and efficiency and stronger slow oscillation-spindle coupling were associated with better future cognitive performance. Importantly, we examined whether these same associations were observed with increased risk of dementia. Though some cohorts demonstrated significant associations between the sleep quantity and efficiency and slow oscillation-spindle coupling composites and dementia risk, pooled estimates across the studies did not show any significant associations, despite similar, oberved directional effects.

A lack of consensus exists regarding whether poor sleep quantity and efficiency are associated with future cognition and dementia risk, with mixed findings to date^24^. A major limitation of many previous studies includes the reliance on self-report sleep, which is subject to misreporting and recall bias, or actigraphy data, which is not able to distinguish sleep stages. In this study, we overcome these past limitations, complementing self-report data with objective data obtained by standardized PSG. We identified a unique cluster of sleep metrics (i.e., PSG and self-report sleep duration, sleep efficiency, wake after sleep onset and non-rapid eye movement sleep cycles) that loaded onto a latent factor reflecting sleep quantity and efficiency. Using this data-driven approach, we show longer and more consolidated sleep was associated with better cognitive performance within 5 years of follow-up. These findings give further weight to previous studies demonstrating that shorter sleep duration^25–27^ and low sleep efficiency^10, 28, 29^ are associated with poorer cognition. Some studies suggest that self-reported longer^25, 30^, rather than short sleep duration, is associated with poorer cognition; however, this study did not support these findings. These differences could be due to how sleep duration was assessed. Self-reported long sleep is reliant on how questions are asked and misclassification can occur due to reporting bed vs sleep time. Importantly, the CLV approach considers sleep duration and efficiency measures in a more global way, rather than in isolation, perhaps pinpointing the sleep trait (i.e, shorter and unconsolidated sleep) that is most important for global cognitive deficits.

Whereas sleep quantity and efficiency were associated with global cognition, this sleep composite was not significantly associated with dementia risk across cohorts. Smaller studies using multi-night actigraphy have identified an association of shorter sleep duration^4^ and lower sleep efficiency^31^ with increased risk of dementia. Therefore, while these individual metrics have been associated with dementia risk, we did not find evidence to suggest that their combination (derived from PSG and self-report sleep measures) was associated with dementia. These inconsistencies may stem from methodological differences in sleep assessment (e.g., multi-night actigraphy vs single night PSG). One possibility is that different sleep characteristics may exert their effects on cognitive health over short time scales. For example, disruptions in sleep continuity may have more immediate effects that are less detectable over the longer follow-up periods used in our dementia analyses. Although assumptions of proportional hazards were met, the extended follow-up may have attenuated time-sensitive associations. Sleep and dementia relationships may also be age dependent. Data from the Whitehall II study of over 6,500 participants shows that self-reported short sleep duration was associated with increased dementia risk only in mid (5^th^ and 6^th^ decades), rather than late life (7^th^ decade)^4^. Despite being the largest study of its kind, we did not have the power to examine whether sleep-dementia relationships varied with age. However, we did observe that the sleep quantity and efficiency composite was significantly associated with reduced dementia risk in the youngest cohort (FHS), but not the older cohorts. It is also possible that individuals with subtle or prodromal cognitive impairment were less likely to participate or complete sleep assessments, potentially attenuating observed associations. Future studies with a larger sample size and range of ages are warranted to address this question.

Since the landmark studies by Xie et al^2^, much attention has been given to the link between dementia and N3 sleep. N3 sleep is thought to be highly important in memory formation^32^ and experimental sleep studies indicate that disruption of N3 sleep increases CSF amyloid-β^33^. Despite these findings, the pooled effects revealed no association between reduced N3 sleep and poorer cognition or dementia risk in this study, though higher N3 sleep composite scores were associated with better cognition in the MrOS cohort. The association between N3 sleep and dementia risk may not be straight forward, as increased amplitude of slow waves may represent heterogenous phenotypes, especially in older individuals, where pathological slowing may confound interpretation of N3 duration. Notably, we did identify that the stronger slow oscillation-spindle coupling during N3 sleep was associated with better cognitive performance in the pooled analysis. Furthermore, stronger slow oscillation-spindle coupling was associated with a non-significant reduction in dementia risk, with this association being significant in the CHS cohort only. EEG analysis in older adults has also identified that stronger slow oscillation-spindle coupling is predictive of better cognitive performance^34^. Importantly, in humans, slow oscillation-spindle coupling is enhanced by prior learning^35^. Stronger slow oscillation-spindle coupling reflects a more consistent and precise alignment of spindles with the up-phases of slow oscillations. It is thought that this precise synchronization and timing increases the efficiency of memory consolidation processes^36^. Our data suggest that additional information on sleep synaptic and oscillatory activity can be gathered by quantifying sleep micro-architecture rather than relying solely on sleep stage distributions.

Increasing evidence also suggests that individual differences in REM sleep may be associated with dementia risk^12, 37^. In this meta-analysis, associations between REM-related composites and brain health were not evident; however, some interesting patterns did emerge. There was a tendency for more and shorter REM sleep bouts to be associated with higher dementia risk in 4 out of 5 cohorts, though the CIs for all crossed 1, indicating non-significance. Further, the sleep composite reflecting less REM% (and more N1/N2 sleep) was associated with poorer global cognitive performance in 2 of the 5 cohorts (CHS and FHS), but this association did not translate to the pooled estimate.

Using SDC data, we have previously identified that OSA of all severities, defined by the apnea hypopnea index (AHI), is associated with poorer cognition^10^. OSA has also been associated with higher risk of dementia in older women^38^. A recent meta-analysis identified a 28% increased risk in clinical Alzheimer’s disease among individuals with OSA^39^. In this study, we identified no association between the respiratory disturbance composite and the outcomes. The respiratory disturbance composite was comprised of AHI, % time spent in oxygen saturation <90%, no. apnea/hypopneas events, and subjective sleepiness. It could be that the metrics that correlate highly to form this composite are not the optimal combination to predict cognitive and dementia outcomes. Emerging research has identified the possible roles of quantitative metrics of hypoxia in predicting adverse outcomes, as well as a possible protective association between respiratory-related arousal and cognition, suggesting the need for a more nuanced approach for understanding the influence of OSA on cognitive outcomes^40^.

Several mechanisms could account for the links between the sleep composites and cognition. Shorter sleep duration is known to increase inflammation^41^ and has been associated with vascular brain injury,^42^ which could potentially promote subsequent cognitive impairment. Furthermore, given that slow oscillation-spindle coupling is thought to be vital for memory formation, the uncoupling of these brain rhythms could disrupt the coordination of information transfer between brain regions (e.g., hippocampal and neocortical) required for the consolidation of memory. In support of this, recent animal studies show that suppression of coordinated patterns of slow oscillation and spindles has disruptive effects on sleep-dependent memory consolidation^43^.

## Strengths and Limitations

This study has a number of strengths, including the use of both self-report and PSG-derived sleep measures, the large sample size, and long-term follow-up of dementia incidence. There are several limitations to this study that should be noted, however. Firstly, while associations were identified between sleep and cognition, we cannot confirm that this relationship is causal in nature. Objective sleep was measured via single-night PSG. While this reflects current clinical protocols, future studies should consider multi-night sleep recordings as more stable estimates of sleep may provide new biological insights. Lastly, our pooled sample was predominantly White, limiting generalizability to other groups.

## Conclusions

This study underscores the role of sleep in supporting optimal cognition, showing that longer, more consolidated sleep and stronger slow oscillation-spindle couplings are associated with better cognitive performance within the next 5 years. However, the lack of consistent associations with dementia risk suggests that any impact of sleep on dementia risk may be more nuanced, potentially influenced by factors not considered here, such as age-dependent sleep biomarkers, genetic factors beyond APOE or any other risk factors that increase vulnerability. By shifting focus from isolated sleep metrics to broader sleep constructs, our study helps to refine our understanding of sleep’s role in cognitive health. Future research could explore whether targeting specific sleep processes—particularly sleep consolidation or slow oscillation-spindle coupling—can enhance cognition and build brain resilience in the face of aging.

## ACKNOWLEDGEMENTS

We thank the participants for dedicating their time to our research. We also thank the researchers involved in data collection and obtaining funding, including Anne Newman (MD, MPH) and John Robbins (MD, MHS) from the Cardiovascular Health Study, who received no compensation for the current article.

## FUNDING

### SDC

The SDC is funded by the NIA (R01 AG062531). This work is also made possible by grants from the NIA to the Cross Cohorts Consortium (CCC) (AG059421) and by the Cohorts for Age and Aging Research in Genomic Epidemiology (CHARGE) infrastructure grant from the NHLBI (HL105756).

### FHS

This work was made possible by grants from the National Institutes of Health (N01-HC-25195, HHSN268201500001I, 75N92019D00031) and the National Institute on Aging (AG059421, AG054076, AG049607, AG033090, AG066524, NS017950, P30AG066546, UF1NS125513).

### ARIC

The ARIC portion of the SHHS was supported by National Heart, Lung, and Blood Institute cooperative agreements U01HL53934 (University of Minnesota) and U01HL64360 (Johns Hopkins University). The Atherosclerosis Risk in Communities study has been funded in whole or in part with Federal funds from the National Heart, Lung, and Blood Institute, National Institutes of Health, Department of Health and Human Services, under Contract nos. (75N92022D00001, 75N92022D00002, 75N92022D00003, 75N92022D00004, 75N92022D00005). The ARIC Neurocognitive Study is supported by U01HL096812, U01HL096814, U01HL096899, U01HL096902, and U01HL096917 from the NIH (NHLBI, NINDS, NIA and NIDCD). The authors thank the staff and participants of the ARIC study for their important contributions.

### CHS

This research was supported by contracts HHSN268201200036C, HHSN268200800007C, HHSN268201800001C, N01HC55222, N01HC85079, N01HC85080, N01HC85081, N01HC85082, N01HC85083, N01HC85086, 75N92021D00006, and grants U01HL080295, U01HL130114 and R01HL172803 from the National Heart, Lung, and Blood Institute (NHLBI), with additional contribution from the National Institute of Neurological Disorders and Stroke (NINDS). Support for cognitive measures was provided by R01AG15928. Additional support was provided by R01AG023629 from the National Institute on Aging (NIA). A full list of principal CHS investigators and institutions can be found at https://CHS-NHLBI.org.

### MrOS

MrOS is supported by National Institutes of Health funding. The following institutes provide support: the National Institute on Aging (NIA), the National Institute of Arthritis and Musculoskeletal and Skin Diseases (NIAMS), the National Center for Advancing Translational Sciences (NCATS), and NIH Roadmap for Medical Research under the following grant numbers: U01 AG027810, U01 AG042124, U01 AG042139, U01 AG042140, U01 AG042143, U01 AG042145, U01 AG042168, U01 AR066160, and UL1 TR002369 The National Heart, Lung, and Blood Institute (NHLBI) provides funding for the MrOS Sleep ancillary study “Outcomes of Sleep Disorders in Older Men” under the following grant numbers: R01 HL071194, R01 HL070848, R01 HL070847, R01 HL070842, R01 HL070841, R01 HL070837, R01 HL070838, and R01 HL070839.

### SOF

SOF is supported by National Institutes of Health funding. The National Institute on Aging (NIA) provides support under the following grant numbers: R01 AG005407, R01 AR35582, R01 AR35583, R01 AR35584, R01 AG005394, R01 AG027574, R01 AG027576, and R01 AG026720.

### Other

Dr. Yiallourou is funded by the American Alzheimer’s Association (AARG-NTF-22-971405). Dr. Pase is supported by the National Health and Medical Research Council of Australia (GTN2009264; GTN1158384). Dr. Cavuoto and Dr. Pase are supported by a Dementia Australia Research Foundation award (Lucas’ Papaw Remedies Project Grant). Dr. Baril is funded by grants from the Fonds de la recherche en santé du Québec, Alzheimer Society of Canada, Canadian Institutes of Health Research, and Sleep Research Society Foundation. Dr. Redline is partially funded by NIA AG 070867. Dr. Yaffe is partially funded by NIA R35 AG071916. Dr. Lutsey is partially supported by K24 HL159246. Dr. Gottesman is supported by the NINDS Intramural Research Program. This research was supported in part by the Intramural Research Program of the National Institutes of Health (NIH). The contributions of the NIH author were made as part of their official duties as NIH federal employees, are in compliance with agency policy requirements, and are considered Works of the United States Government. However, the findings and conclusions presented in this paper are those of the author and do not necessarily reflect the views of the NIH or the U.S. Department of Health and Human Services. Dr. Purcell is partially funded by NIH NHLBI R01HL146339, NIH NIA 070867, and NIH NIMHD MD012738. Dr. Leng is partially funded by NIA R01AG083836. Drs. Seshadri and Himali are partially supported by the South Texas Alzheimer’s Disease Center (1P30AG066546-01A1) and The Bill and Rebecca Reed Endowment for Precision Therapies and Palliative Care. Dr. Seshadri is also supported by an endowment from the Barker Foundation as the Robert R Barker Distinguished University Professor of Neurology, Psychiatry and Cellular and Integrative Physiology and Dr. Himali by an endowment from the William Castella family as William Castella Distinguished University Chair for Alzheimer’s Disease Research. Drs. Redline and Purcell were partially supported by 5R01AG070867 and NIH contract 75N92019C00011. Mr. Weihs is supported by the German Research Foundation (DFG, Grant number: 406711066).

### Financial Disclosures

Katie Stone received grant funding from Eli Lilly (related to screening for OSA in primary care) and was Consultant on DSMB for Axsome Therapeutics (for studies in their Somriamfetol portfolio). Professor Susan Redline has received consulting fees from Amgen Inc, grant funding from Google, is an unpaid scientific advisor to Apnimed, and is an unpaid Board member for the Alliance of Sleep Apnea Partners. She receives a stipend from the National Sleep Foundation for her role as editor of Sleep Health.

