## Supplement for "A data driven approach to assess relationships between sleep, cognition and dementia: Findings from the Sleep and Dementia Consortium"

**Supplementary Tables:**

**eTable 1: Timing of Assessments**

|  | PSG dates (years) | Cognitive dates (years) | Dementia follow-dates (years)* |
| --- | --- | --- | --- |
| ARIC | 1996-1998 | 1996-1998 | 1996-2020 |
| CHS | 1995-1998 | 1999-2000 | 1995 - 06/2013 |
| FHS | 1995-1998 | 1999-2002 | 1995 - 12/31/2018 |
| MrOS | 2003-2005 | 2003-2005 | 2009-2012 |
| SOF | 2002-2004 | 2006-2008 | 2006-2008 |

*ARIC and FHS both had continuous surveillance of dementia with dementia adjudicated by a committee according to DSM-IV/V criteria (or equivalent). CHS also adjudicated dementia based on DSM-IV criteria. For ARIC, cases were identified by hospitalization and death ICD-9 codes until 2011-2013 when intensive in-clinic cognitive testing and phone surveillance began. Both MrOS and SOF assessed dementia at discrete follow-up time points (e.g., several years), up to approximately 10- and 5-years following sleep assessment, respectively.

**eTable 2: Sample selection**

| **Cognitive analysis sample** | | | | | |
| --- | --- | --- | --- | --- | --- |
| Cohort | ARIC | CHS | FHS | MrOS | SOF |
| Participants with PSG and NP | 1879 | 836 | 756 | 2799 | 240 |
| Participants with <180 minutes of Total Sleep Time or <1 minute of REM sleep | 48 | 43 | 13 | 77 | 4 |
| Younger than 45 at time of PSG | 0 | 0 | 66 | 0 | 0 |
| Prevalent dementia | 3 | 21 | 6 | 0 | 0 |
| Prevalent stroke (or missing prevalent stroke) | 34 | 25 | 17 | 103 | 44 |
| Missing covariates | 3 | 35 | 10 | 0 | 0 |
| Final cognitive sample | 1791 | 712 | 644 | 2619 | 192 |
| **Dementia analysis sample (model 1)** | | | | | |
| Cohort | ARIC | CHS | FHS | MrOS | SOF |
| Participants with PSG | 1915 | 1229 | 920 | 2911 | 461 |
| Participants with <180 minutes of Total Sleep Time or <1 minute of REM sleep | 50 | 82 | 1 | 95 | 21 |
| Younger than 60 at time of PSG | 680 | 0 | 541 | 0 | 0 |
| Prevalent dementia | 3 | 81 | 0 | 140 | 4 |
| Other neurological condition(s) | 28 | 72 | 8 | 96 | 58 |
| Missing covariates | 1 | 4 | 0 | 0 | 0 |
| Final dementia sample | 1153 | 990 | 370 | 2580 | 378 |

| **eTable 3. Cognitive tests for global cognitive composite scores** | |
| --- | --- |
| Cohort  Cognitive task^a^ | Component loading^b^ |
| **ARIC** |  |
| Delayed Word Recall Test, N correct | 0.38 |
| Digit Symbol Substitution Test, N correct | 0.48 |
| Verbal Fluency, N correct | 0.45 |
| **CHS** |  |
| Modified MMSE (3MS), N correct | 0.69 |
| Digit symbol substitution test, N correct | 0.79 |
| Benton visual retention test, N correct | 0.67 |
| Trails A, sec | 0.67 |
| Trails B, sec | 0.81 |
| **FHS** |  |
| Trails B, sec | 0.35 |
| Logical Memory^b^, N correct | 0.31 |
| Visual Reproductions^c^, N correct | 0.37 |
| Similarities, N correct | 0.35 |
| **MrOS** |  |
| Trails B, sec | 0.41 |
| Digit Vigilance, sec | 0.48 |
| Modified MMSE (3MS), N correct | 0.41 |
| **SOF** |  |
| Digital span Forward, N correct | 0.18 |
| California Verbal Learning Test short form, N correct | 0.45 |
| Trails B, sec | 0.40 |
| Category Fluency Task (vegetables over one minute interval, N correct | 0.41 |
| ^a^Transformed cognitive tasks from the Atherosclerosis Risk in Communities (ARIC) study, Cardiovascular Health Study (CHS), Framingham Heart Study (FHS), Osteoporotic Fractures in Men Study (MrOS) and Study of Osteoporotic Fractures (SOF) were used to create the standardized variables, where applicable.  ^b^The overall global cognitive score was calculated by summing the products of the standardizing formulas and the component loadings for each cognitive task.  ^c^Immediate and Delayed recall scores were summed together into a single variable | |

**eTable 4: Cohort characteristics at the time of the sleep study – dementia sample**

|  | ARIC | CHS | FHS | MrOS | SOF |
| --- | --- | --- | --- | --- | --- |
| Participants who meet the inclusion criteria | 1153 | 990 | 370 | 2580 | 378 |
| Age, years | 65.9 (3.9) | 77.3 (4.3) | 67.5 (4.9) | 76.1 (5.4) | 82.8 (3.4) |
| 60-70 | 905 (78.5) | 12 (1.2) | 253 (68.4) | 241 (9.3) | 0 (0.0) |
| 70-80 | 248 (21.5) | 710 (71.7) | 114 (30.8) | 902 (35.0) | 41 (10.9) |
| 80-90 | 0 (0.0) | 254 (25.7) | 3 (0.8) | 754 (29.2) | 320 (84.7) |
| 90+ | 0 (0.0) | 14 (1.4) | 0 (0.0) | 683 (26.5) | 17 (4.5) |
| Female, n(%) | 555 (48.1) | 562 (56.8) | 187 (50.5) | 0 (0.0) | 378 (100.0) |
| Race | | | | | |
| White, n (%) | 1148 (99.6) | 821 (82.9) | 317 (85.7) | 2359 (91.4) | 349 (92.3) |
| Black, n (%) | 3 (0.3) | 165 (16.7) | 32 (8.7) | 73 (2.8) | 28 (7.4) |
| Other, n (%) | 2 (0.2) | 4 (0.4) | 21 (5.7) | 148 (5.7) | 1 (0.3) |
| Education | | | | | |
| <12 years high school, n (%) | 159 (13.8) | 204 (20.6) | 39 (10.8) | 108 (4.2) | 67 (19.1) |
| 12 years high school , n (%) | 420 (36.4) | 537 (54.3) | 116 (32.0) | 408 (15.8) | 167 (47.7) |
| >12 years high school, n (%) | 572 (49.6) | 248 (25.1) | 207 (57.2) | 2064 (80.0) | 116 (33.1) |
| Systolic blood pressure, mmHg | 123.78 (17.8) | 130.35 (19.4) | 131.3 (18.3) | 126.64 (16.0) | 135.96 (16.7) |
| Treatment for hypertension, n (%) | 452 (39.2) | 569 (57.5) | 134 (36.4) | 1703 (66.0) | 272 (72.0) |
| Stage I Hypertension, n(%) | 467 (40.5) | 586 (59.2) | 190 (51.8) | 1838 (71.2) | 311 (82.7) |
| Prevalent diabetes, n (%) | 161 (14.0) | 130 (13.2) | 47 (12.8) | 339 (13.1) | 43 (11.4) |
| Prevalent CVD, n (%) | 220 (19.1) | 116 (11.7) | 41 (11.1) | 1015 (39.4) | 78 (20.6) |
| Smoking status [current or not], n (%) | 104 (9.0) | 57 (5.8) | 39 (10.5) | 51 (2.0) | 9 (2.4) |
| Body mass index, Median [Q1, Q3] | 28.2 [25.3, 31.8] | 27.3 [24.5, 29.9] | 27 [25, 31] | 26.8 [24.6, 29.3] | 27.3 [24.5, 31.1] |
| Regular use of sleep pills, n (%) | 271 (23.5) | 223 (22.9) | 50 (13.9) | 522 (20.2) | 95 (25.1) |
| Use of antidepressants, n (%) | 73 (6.3) | 54 (5.5) | 10 (2.7) | 178 (6.9) | 35 (9.2) |
| Use of sedatives, n (%) | 59 (5.1) | 75 (7.6) | 13 (3.5) | 174 (6.7) | 57 (15.1) |
| Positivity for an APOE ε4 allele, n (%) | 309 (26.8) | 223 (23.7) | 81 (22.3) | 484 (23.2) | 22 (10.9) |

Abbreviations: ARIC, Atherosclerosis Risk in Communities study; CHS, Cardiovascular Health Study; CVD, cardiovascular disease; FHS, Framingham Heart Study; MrOS, Osteoporotic Fractures in Men Study; SOF, Study of Osteoporotic Fractures.

eTable 5. **Associations (β (95% CIs) p-value) between sleep composites and cognitive outcomes**

| **Sleep Composite** | **ARIC** | **CHS** | **FHS** | **MrOS** | **SOF** | **All Cohorts** |
| --- | --- | --- | --- | --- | --- | --- |
| **Global Cognition** | | | | | |  |
| Sleep quantity and efficiency | 0.007 (-0.019, 0.033) 0.599 | 0.037 (-0.003, 0.077) 0.068 | 0.036 (-0.006, 0.078) 0.089 | 0.050 (0.024, 0.076) <.001 | 0.026 (-0.076, 0.129) 0.611 | 0.031 (0.004; 0.058)  0.033 |
| Sleep fragmentation | 0.007 (-0.015, 0.029) 0.535 | 0.013 (-0.023, 0.049) 0.477 | 0.007 (-0.030, 0.043) 0.716 | 0.004 (-0.018, 0.027) 0.701 | -0.033 (-0.118, 0.052) 0.447 | 0.006 (-0.016; 0.028)  0.496 |
| Light NREM predominance | -0.020 (-0.051, 0.011) 0.211 | -0.052 (-0.101, -0.003) 0.038 | -0.054 (-0.103, -0.006) 0.028 | -0.016 (-0.049, 0.017) 0.339 | 0.049 (-0.058, 0.155) 0.366 | -0.027 (-0.072; 0.018)  0.173 |
| N3 predominance | -0.007 (-0.027, 0.014) 0.521 | -0.006 (-0.036, 0.025) 0.716 | 0.004 (-0.030, 0.037) 0.824 | 0.028 (0.002, 0.053) 0.033 | -0.006 (-0.086, 0.075) 0.886 | 0.004 (-0.019; 0.027)  0.641 |
| Spindle number and duration | -0.001 (-0.021, 0.018) 0.897 | -0.019 (-0.053, 0.015) 0.277 | 0.031 (-0.0001, 0.061) 0.050 | -0.007 (-0.030, 0.016) 0.564 | -0.012 (-0.090, 0.065) 0.757 | -0.000 (-0.026; 0.026)  0.999 |
| REM sleep bouts | 0.002 (-0.030, 0.033) 0.921 | -0.008 (-0.056, 0.040) 0.752 | -0.045 (-0.098, 0.009) 0.102 | 0.005 (-0.021, 0.032) 0.699 | 0.003 (-0.116, 0.121) 0.967 | -0.005 (-0.036; 0.026)  0.681 |
| Respiratory disturbances | 0.008 (-0.015, 0.032) 0.480 | 0.001 (-0.041, 0.044) 0.949 | 0.028 (-0.008, 0.064) 0.130 | -0.003 (-0.023, 0.017) 0.789 | 0.076 (-0.022, 0.173) 0.128 | 0.011 (-0.022; 0.044)  0.402 |
| Slow oscillation-spindle coupling | 0.030 (0.001, 0.058) 0.041 | 0.062 (0.011, 0.112) 0.016 | 0.043 (-0.004, 0.091) 0.075 | 0.020 (-0.012, 0.052) 0.225 | 0.089 (-0.030, 0.209) 0.143 | 0.037 (0.003; 0.070)  0.039 |
| Spindle amplitude | 0.001 (-0.026, 0.029) 0.916 | 0.005 (-0.038, 0.049) 0.806 | -0.029 (-0.075, 0.017) 0.220 | -0.010 (-0.036, 0.016) 0.444 | -0.077 (-0.194, 0.041) 0.199 | -0.010 (-0.045; 0.025)  0.479 |
| **Attention and processing speed** | | | | | | |
| Sleep quantity and efficiency | 0.010 (-0.012, 0.031) 0.368 | 0.035 (-0.021, 0.090) 0.219 | -0.009 (-0.057, 0.040) 0.728 | 0.021 (-0.007, 0.049) 0.136 | 0.016 (-0.084, 0.116) 0.750 | 0.054 (-0.086; 0.194)  0.344 |
| Sleep fragmentation | -0.003 (-0.021, 0.016) 0.759 | 0.011 (-0.040, 0.062) 0.676 | 0.014 (-0.029, 0.056) 0.521 | 0.0003 (-0.024, 0.025) 0.979 | -0.001 (-0.083, 0.082) 0.989 | -0.013 (-0.067; 0.042)  0.548 |
| Light NREM predominance | -0.025 (-0.050, 0.001) 0.055 | -0.068 (-0.140, 0.003) 0.059 | -0.038 (-0.094, 0.019) 0.189 | -0.020 (-0.055, 0.015) 0.263 | 0.127 (0.025, 0.229) 0.015 | -0.017 (-0.106; 0.073)  0.635 |
| N3 predominance | 0.0004 (-0.016, 0.017) 0.959 | 0.014 (-0.032, 0.060) 0.554 | 0.013 (-0.025, 0.052) 0.499 | 0.017 (-0.010, 0.044) 0.223 | -0.002 (-0.079, 0.075) 0.951 | 0.007 (-0.061; 0.075)  0.787 |
| Spindle number and duration | 0.001 (-0.015, 0.017) 0.914 | -0.052 (-0.102, -0.001) 0.045 | -0.003 (-0.039, 0.033) 0.869 | 0.001 (-0.024, 0.026) 0.944 | 0.006 (-0.069, 0.081) 0.875 | -0.015 (-0.075; 0.046)  0.538 |
| REM sleep bouts | -0.010 (-0.036, 0.016) 0.441 | -0.001 (-0.074, 0.072) 0.980 | -0.017 (-0.079, 0.045) 0.585 | 0.005 (-0.024, 0.033) 0.749 | 0.103 (-0.011, 0.216) 0.076 | -0.006 (-0.096; 0.084)  0.861 |
| Respiratory disturbances | 0.013 (-0.006, 0.032) 0.172 | 0.011 (-0.054, 0.076) 0.732 | 0.013 (-0.030, 0.056) 0.558 | -0.007 (-0.028, 0.014) 0.522 | 0.079 (-0.015, 0.173) 0.101 | 0.085 (-0.305; 0.474)  0.579 |
| Slow oscillation-spindle coupling | 0.023 (-0.001, 0.046) 0.056 | 0.025 (-0.048, 0.098) 0.493 | -0.006 (-0.061, 0.050) 0.843 | 0.008 (-0.026, 0.043) 0.634 | 0.103 (-0.012, 0.219) 0.080 | 0.004 (-0.245; 0.254)  0.963 |
| Spindle amplitude | -0.004 (-0.027, 0.019) 0.738 | -0.016 (-0.078, 0.046) 0.608 | 0.001 (-0.053, 0.055) 0.971 | -0.016 (-0.044, 0.012) 0.274 | 0.089 (-0.025, 0.202) 0.124 | 0.009 (-0.074; 0.092)  0.787 |
| **Executive Function** | | | | | |  |
| Sleep quantity and efficiency | -0.011 (-0.038, 0.017) 0.446 | 0.037 (-0.012, 0.087) 0.136 | 0.021 (-0.014, 0.057) 0.241 | 0.042 (0.015, 0.069) 0.002 | -0.013 (-0.113, 0.088) 0.806 | 0.019 (-0.015; 0.053)  0.197 |
| Sleep fragmentation | 0.031 (0.007, 0.054) 0.010 | 0.007 (-0.039, 0.052) 0.776 | -0.008 (-0.039, 0.023) 0.627 | 0.004 (-0.019, 0.027) 0.728 | 0.004 (-0.079, 0.088) 0.917 | 0.010 (-0.014; 0.034)  0.300 |
| Light NREM predominance | 0.003 (-0.030, 0.036) 0.842 | -0.039 (-0.102, 0.024) 0.220 | -0.031 (-0.073, 0.010) 0.137 | -0.025 (-0.058, 0.009) 0.148 | -0.0002 (-0.104, 0.104) 0.997 | -0.018 (-0.049; 0.013)  0.185 |
| N3 predominance | -0.017 (-0.038, 0.004) 0.114 | 0.034 (-0.004, 0.073) 0.082 | 0.003 (-0.026, 0.031) 0.860 | 0.026 (-0.0002, 0.052) 0.051 | -0.024 (-0.103, 0.056) 0.556 | 0.007 (-0.025; 0.038)  0.589 |
| Spindle number and duration | -0.001 (-0.022, 0.020) 0.942 | -0.042 (-0.085, 0.002) 0.059 | 0.012 (-0.014, 0.038) 0.368 | -0.015 (-0.038, 0.009) 0.226 | -0.014 (-0.090, 0.062) 0.711 | -0.008 (-0.034; 0.019)  0.474 |
| REM sleep bouts | 0.005 (-0.029, 0.038) 0.775 | -0.021 (-0.084, 0.042) 0.512 | -0.035 (-0.081, 0.010) 0.125 | -0.001 (-0.028, 0.026) 0.916 | 0.052 (-0.064, 0.168) 0.378 | -0.007 (-0.044; 0.031)  0.647 |
| Respiratory disturbances | 0.020 (-0.004, 0.045) 0.099 | 0.007 (-0.048, 0.062) 0.800 | 0.010 (-0.021, 0.042) 0.512 | -0.0004 (-0.021, 0.020) 0.968 | 0.060 (-0.036, 0.156) 0.219 | 0.011 (-0.015; 0.037)  0.305 |
| Slow oscillation-spindle coupling | 0.023 (-0.006, 0.053) 0.124 | 0.024 (-0.040, 0.088) 0.460 | 0.026 (-0.014, 0.066) 0.205 | 0.026 (-0.007, 0.059) 0.120 | 0.011 (-0.107, 0.128) 0.859 | 0.025 (-0.001; 0.050)  0.059 |
| Spindle amplitude | -0.011 (-0.041, 0.018) 0.453 | -0.025 (-0.079, 0.028) 0.355 | -0.018 (-0.058, 0.021) 0.360 | -0.006 (-0.032, 0.021) 0.686 | -0.034 (-0.150, 0.081) 0.559 | -0.012 (-0.036; 0.012)  0.232 |
| **Language** | | | | | |  |
| Sleep quantity and efficiency |  | -0.005 (-0.064, 0.054) 0.871 | -0.00007 (-0.045, 0.045) 0.998 |  | -0.006 (-0.108, 0.096) 0.902 | -0.002 (-0.076; 0.072)  0.904 |
| Sleep fragmentation |  | 0.013 (-0.042, 0.069) 0.633 | 0.005 (-0.035, 0.045) 0.808 |  | -0.027 (-0.111, 0.057) 0.525 | 0.003 (-0.068; 0.074)  0.871 |
| Light NREM predominance |  | -0.047 (-0.123, 0.030) 0.229 | 0.016 (-0.037, 0.068) 0.562 |  | 0.025 (-0.080, 0.131) 0.638 | -0.002 (-0.112; 0.108)  0.944 |
| N3 predominance |  | 0.012 (-0.036, 0.061) 0.619 | -0.023 (-0.059, 0.013) 0.215 |  | 0.0002 (-0.079, 0.079) 0.997 | -0.008 (-0.074; 0.058)  0.647 |
| Spindle number and duration |  | -0.003 (-0.057, 0.051) 0.918 | 0.010 (-0.023, 0.043) 0.568 |  | -0.034 (-0.111, 0.042) 0.379 | -0.000 (-0.067; 0.067)  0.993 |
| REM sleep bouts |  | 0.041 (-0.036, 0.119) 0.292 | -0.062 (-0.120, -0.005) 0.032 |  | -0.046 (-0.163, 0.072) 0.444 | -0.023 (-0.170; 0.124)  0.574 |
| Respiratory disturbances |  | 0.008 (-0.060, 0.077) 0.809 | 0.021 (-0.020, 0.062) 0.320 |  | 0.046 (-0.052, 0.143) 0.355 | 0.021 (-0.054; 0.096)  0.354 |
| Slow oscillation-spindle coupling |  | -0.019 (-0.095, 0.057) 0.622 | 0.031 (-0.021, 0.082) 0.241 |  | 0.084 (-0.034, 0.203) 0.163 | 0.024 (-0.103; 0.151)  0.504 |
| Spindle amplitude |  | -0.015 (-0.081, 0.050) 0.648 | -0.026 (-0.076, 0.024) 0.308 |  | -0.096 (-0.212, 0.020) 0.103 | -0.033 (-0.141; 0.075)  0.322 |
| **Verbal Learning and Memory** | | | | | |  |
| Sleep quantity and efficiency | 0.019 (-0.027, 0.065) 0.424 | 0.009 (-0.064, 0.081) 0.815 | 0.015 (-0.025, 0.054) 0.473 |  | 0.069 (-0.038, 0.177) 0.205 | 0.019 (-0.030; 0.068)  0.304 |
| Sleep fragmentation | -0.014 (-0.054, 0.025) 0.483 | 0.018 (-0.050, 0.087) 0.595 | 0.008 (-0.027, 0.043) 0.646 |  | -0.052 (-0.142, 0.037) 0.250 | -0.004 (-0.054; 0.046)  0.813 |
| Light NREM predominance | -0.025 (-0.080, 0.030) 0.376 | -0.067 (-0.159, 0.026) 0.156 | -0.033 (-0.080, 0.013) 0.159 |  | 0.035 (-0.079, 0.148) 0.547 | -0.028 (-0.095; 0.039)  0.276 |
| N3 predominance | 0.002 (-0.033, 0.038) 0.900 | -0.023 (-0.082, 0.037) 0.449 | 0.009 (-0.023, 0.041) 0.588 |  | 0.010 (-0.076, 0.095) 0.825 | 0.002 (-0.035; 0.039)  0.864 |
| Spindle number and duration | -0.004 (-0.038, 0.031) 0.837 | 0.010 (-0.056, 0.077) 0.764 | 0.020 (-0.009, 0.049) 0.179 |  | 0.014 (-0.067, 0.096) 0.729 | 0.010 (-0.024; 0.045)  0.410 |
| REM sleep bouts | 0.011 (-0.045, 0.067) 0.691 | -0.018 (-0.113, 0.078) 0.717 | -0.064 (-0.114, -0.013) 0.013 |  | -0.043 (-0.167, 0.082) 0.500 | -0.028 (-0.096; 0.040)  0.280 |
| Respiratory disturbances | -0.019 (-0.060, 0.023) 0.376 | 0.013 (-0.075, 0.100) 0.776 | 0.029 (-0.006, 0.064) 0.106 |  | 0.040 (-0.063, 0.144) 0.444 | 0.011 (-0.040; 0.062)  0.550 |
| Slow oscillation-spindle coupling | 0.021 (-0.028, 0.071) 0.397 | 0.031 (-0.063, 0.125) 0.515 | 0.010 (-0.035, 0.056) 0.651 |  | 0.071 (-0.055, 0.198) 0.266 | 0.021 (-0.033; 0.075)  0.301 |
| Spindle amplitude | 0.022 (-0.027, 0.071) 0.376 | -0.016 (-0.097, 0.065) 0.699 | -0.008 (-0.052, 0.036) 0.731 |  | -0.088 (-0.212, 0.035) 0.161 | -0.008 (-0.078; 0.063)  0.758 |
| **Visuospatial Function** | | | | | |  |
| Sleep quantity and efficiency |  | 0.003 (-0.059, 0.065) 0.921 | 0.021 (-0.019, 0.062) 0.295 |  |  | 0.016 (-0.207; 0.239)  0.530 |
| Sleep fragmentation |  | 0.020 (-0.036, 0.076) 0.475 | 0.022 (-0.013, 0.058) 0.216 |  |  | 0.022 (-0.172; 0.215)  0.390 |
| Light NREM predominance |  | -0.101 (-0.180, -0.022) 0.012 | -0.050 (-0.097, -0.004) 0.035 |  |  | -0.067 (-0.403; 0.269)  0.239 |
| N3 predominance |  | -0.022 (-0.072, 0.028) 0.394 | -0.002 (-0.034, 0.030) 0.908 |  |  | -0.008 (-0.193; 0.177)  0.681 |
| Spindle number and duration |  | -0.076 (-0.132, -0.020) 0.008 | 0.036 (0.006, 0.065) 0.017 |  |  | -0.017 (-0.726; 0.692)  0.810 |
| REM sleep bouts |  | -0.006 (-0.087, 0.074) 0.875 | 0.022 (-0.029, 0.073) 0.403 |  |  | 0.013 (-0.277; 0.304)  0.662 |
| Respiratory disturbances |  | -0.062 (-0.134, 0.010) 0.091 | 0.022 (-0.013, 0.058) 0.218 |  |  | -0.013 (-0.543; 0.516)  0.804 |
| Slow oscillation-spindle coupling |  | 0.030 (-0.051, 0.111) 0.462 | 0.073 (0.028, 0.119) 0.002 |  |  | 0.061 (-0.244; 0.365)  0.240 |
| Spindle amplitude |  | -0.018 (-0.088, 0.053) 0.621 | -0.042 (-0.087, 0.002) 0.062 |  |  | -0.035 (-0.288; 0.217)  0.327 |

The directionality for each sleep composite was interpreted as: Sleep quantity = higher with longer sleep; Sleep fragmentation = higher with more fragmented sleep; Light NREM predominance = higher with more N1/N2 events and less REM/N3 events; N3 predominance = higher with more and longer N3; Spindle number and duration = higher value with lower number/ duration of spindles; REM sleep bouts = higher with more and shorter bouts of REM sleep; Respiratory disturbances = higher with more OSA); Spindle and slow wave coupling = higher with stronger coupling; Spindle amplitude = higher with higher amplitude.

Abbreviations: ARIC, Atherosclerosis Risk in Communities study; CHS, Cardiovascular Health Study; FHS, Framingham Heart Study; MrOS, Osteoporotic Fractures in Men Study; SOF, Study of Osteoporotic Fractures; NREM, non-rapid eye movement sleep; N3, non-rapid eye movement sleep; REM, rapid eye movement sleep.

eTable 6. Interaction p-values for sleep composite x APOE ε4 for cognitive outcomes

| **Sleep Composite** | **ARIC** | **CHS** | **FHS** | **MrOS** | **SOF** | **All Cohorts** |
| --- | --- | --- | --- | --- | --- | --- |
| **Global Cognition** | | | | | | |
| Sleep quantity and efficiency | 0.369 | 0.669 | 0.359 | 0.441 | 0.371 | 0.756 |
| Sleep fragmentation | 0.514 | 0.237 | 0.729 | 0.335 | 0.985 | 0.667 |
| Light NREM predominance | 0.500 | 0.476 | 0.321 | 0.505 | 0.228 | 0.891 |
| N3 predominance | 0.709 | 0.689 | 0.660 | 0.230 | 0.720 | 0.722 |
| Spindle number and duration | 0.577 | 0.813 | 0.468 | 0.728 | 0.586 | 0.607 |
| REM sleep bouts | 0.397 | 0.510 | 0.847 | 0.675 | 0.773 | 0.441 |
| Respiratory disturbances | 0.467 | 0.060 | 0.809 | 0.302 | 0.833 | 0.898 |
| Slow oscillation-spindle coupling | 0.777 | 0.901 | 0.604 | 0.654 | 0.294 | 0.756 |
| Spindle amplitude | 0.477 | 0.160 | 0.091 | 0.472 | 0.813 | 0.958 |
| **Attention & Processing Speed** | | | | | | |
| Sleep quantity and efficiency | 0.581 | 0.310 | 0.199 | 0.099 | 0.149 | 0.344 |
| Sleep fragmentation | 0.904 | 0.842 | 0.457 | 0.411 | 0.623 | 0.548 |
| Light NREM predominance | 0.861 | 0.819 | 0.066 | 0.751 | 0.843 | 0.635 |
| N3 predominance | 0.474 | 0.252 | 0.792 | 0.362 | 0.437 | 0.787 |
| Spindle number and duration | 0.845 | 0.101 | 0.705 | 0.766 | 0.809 | 0.538 |
| REM sleep bouts | 0.608 | 0.551 | 0.164 | 0.540 | 0.717 | 0.861 |
| Respiratory disturbances | 0.552 | 0.175 | 0.445 | 0.075 | 0.011 | 0.579 |
| Slow oscillation-spindle coupling | 0.168 | 0.317 | 0.619 | 0.162 | 0.265 | 0.963 |
| Spindle amplitude | 0.327 | 0.993 | 0.403 | 0.510 | 0.442 | 0.787 |
| **Executive Function** | | | | | | |
| Sleep quantity and efficiency | 0.993 | 0.329 | 0.817 | 0.738 | 0.971 | 0.583 |
| Sleep fragmentation | 0.302 | 0.400 | 0.409 | 0.732 | 0.469 | 0.792 |
| Light NREM predominance | 0.716 | 0.976 | 0.149 | 0.355 | 0.066 | 0.960 |
| N3 predominance | 0.140 | 0.057 | 0.417 | 0.164 | 0.937 | 0.405 |
| Spindle number and duration | 0.915 | 0.515 | 0.539 | 0.279 | 0.493 | 0.671 |
| REM sleep bouts | 0.137 | 0.854 | 0.534 | 0.090 | 0.468 | 0.510 |
| Respiratory disturbances | 0.048 | 0.589 | 0.885 | 0.739 | 0.884 | 0.253 |
| Slow oscillation-spindle coupling | 0.291 | 0.367 | 0.668 | 0.235 | 0.585 | 0.962 |
| Spindle amplitude | 0.372 | 0.294 | 0.081 | 0.392 | 0.348 | 0.981 |
| **Language** | | | | | | |
| Sleep quantity and efficiency |  | 0.166 | 0.671 |  | 0.207 | 0.574 |
| Sleep fragmentation |  | 0.298 | 0.556 |  | 0.625 | 0.798 |
| Light NREM predominance |  | 0.853 | 0.027 |  | 0.549 | 0.571 |
| N3 predominance |  | 0.679 | 0.167 |  | 0.368 | 0.693 |
| Spindle number and duration |  | 0.356 | 0.798 |  | 0.366 | 0.637 |
| REM sleep bouts |  | 0.474 | 0.651 |  | 0.425 | 0.783 |
| Respiratory disturbances |  | 0.060 | 0.667 |  | 0.872 | 0.579 |
| Slow oscillation-spindle coupling |  | 0.269 | 0.960 |  | 0.428 | 0.647 |
| Spindle amplitude |  | 0.967 | 0.592 |  | 0.705 | 0.683 |
| **Verbal Learning and Memory** | | | | | | |
| Sleep quantity and efficiency | 0.309 | 0.536 | 0.105 |  | 0.860 | 0.157 |
| Sleep fragmentation | 0.752 | 0.213 | 0.243 |  | 0.717 | 0.919 |
| Light NREM predominance | 0.401 | 0.575 | 0.438 |  | 0.641 | 0.367 |
| N3 predominance | 0.380 | 0.040 | 0.372 |  | 0.775 | 0.928 |
| Spindle number and duration | 0.388 | 0.713 | 0.339 |  | 0.443 | 0.400 |
| REM sleep bouts | 0.609 | 0.477 | 0.752 |  | 0.574 | 0.675 |
| Respiratory disturbances | 0.036 | 0.198 | 0.710 |  | 0.612 | 0.295 |
| Slow oscillation-spindle coupling | 0.716 | 0.391 | 0.477 |  | 0.545 | 0.732 |
| Spindle amplitude | 0.894 | 0.626 | 0.565 |  | 0.845 | 0.883 |
| **Visuospatial Function** | | | | | | |
| Sleep quantity and efficiency |  | 0.261 | 0.342 |  |  | 0.395 |
| Sleep fragmentation |  | 0.370 | 0.806 |  |  | 0.613 |
| Light NREM predominance |  | 0.858 | 0.973 |  |  | 0.929 |
| N3 predominance |  | 0.115 | 0.408 |  |  | 0.377 |
| Spindle number and duration |  | 0.698 | 0.928 |  |  | 0.927 |
| REM sleep bouts |  | 0.285 | 0.511 |  |  | 0.468 |
| Respiratory disturbances |  | 0.502 | 0.413 |  |  | 0.871 |
| Slow oscillation-spindle coupling |  | 0.262 | 0.849 |  |  | 0.725 |
| Spindle amplitude |  | 0.660 | 0.292 |  |  | 0.708 |

The directionality for each sleep composite was interpreted as: Sleep quantity = higher with longer sleep; Sleep fragmentation = higher with more fragmented sleep; Light NREM predominance = higher with more N1/N2 events and less REM/N3 events; N3 predominance = higher with more and longer N3; Spindle number and duration = higher value with lower number/ duration of spindles; REM sleep bouts = higher with more and shorter bouts of REM sleep; Respiratory disturbances = higher with more OSA); Spindle and slow wave coupling = higher with stronger coupling; Spindle amplitude = higher with higher amplitude.

Abbreviations: ARIC, Atherosclerosis Risk in Communities study; CHS, Cardiovascular Health Study; FHS, Framingham Heart Study; MrOS, Osteoporotic Fractures in Men Study; SOF, Study of Osteoporotic Fractures; NREM, non-rapid eye movement sleep; N3, non-rapid eye movement sleep; REM, rapid eye movement sleep.

eTable 7. Interaction p-values for sleep composite x sex for cognitive outcomes

| **Sleep Composite** | **ARIC** | **CHS** | **FHS** | **All Cohorts** |
| --- | --- | --- | --- | --- |
| **Global Cognition** | | | | |
| Sleep quantity and efficiency | 0.445 | 0.352 | 0.155 | 0.936 |
| Sleep fragmentation | 0.803 | 0.538 | 0.437 | 0.493 |
| Light NREM predominance | 0.661 | 0.986 | 0.125 | 0.697 |
| N3 predominance | 0.421 | 0.286 | 0.695 | 0.325 |
| Spindle number and duration | 0.757 | 0.090 | 0.959 | 0.490 |
| REM sleep bouts | 0.615 | 0.665 | 0.937 | 0.597 |
| Respiratory disturbances | 0.130 | 0.444 | 0.974 | 0.701 |
| Slow oscillation-spindle coupling | 0.466 | 0.190 | 0.508 | 0.969 |
| Spindle amplitude | 0.581 | 0.545 | 0.955 | 0.542 |
| **Attention & Processing Speed** | | | | |
| Sleep quantity and efficiency | 0.182 | 0.761 | 0.278 | 0.810 |
| Sleep fragmentation | 0.890 | 0.346 | 0.521 | 0.669 |
| Light NREM predominance | 0.524 | 0.469 | 0.350 | 0.382 |
| N3 predominance | 0.705 | 0.903 | 0.930 | 0.768 |
| Spindle number and duration | 0.516 | 0.595 | 0.415 | 0.411 |
| REM sleep bouts | 0.668 | 0.204 | 0.192 | 0.341 |
| Respiratory disturbances | 0.211 | 0.083 | 0.799 | 0.715 |
| Slow oscillation-spindle coupling | 0.250 | 0.892 | 0.779 | 0.365 |
| Spindle amplitude | 0.727 | 0.185 | 0.622 | 0.695 |
| **Executive functioning** | | | | |
| Sleep quantity and efficiency | 0.635 | 0.701 | 0.036 | 0.484 |
| Sleep fragmentation | 0.901 | 0.826 | 0.790 | 0.890 |
| Light NREM predominance | 0.856 | 0.952 | 0.295 | 0.705 |
| N3 predominance | 0.951 | 0.891 | 0.316 | 0.639 |
| Spindle number and duration | 0.565 | 0.609 | 0.542 | 0.637 |
| REM sleep bouts | 0.809 | 0.701 | 0.797 | 0.685 |
| Respiratory disturbances | 0.414 | 0.337 | 0.191 | 0.646 |
| Slow oscillation-spindle coupling | 0.702 | 0.366 | 0.200 | 0.728 |
| Spindle amplitude | 0.410 | 0.516 | 0.613 | 0.659 |
| **Language** | | | | |
| Sleep quantity and efficiency |  | 0.864 | 0.985 | 0.945 |
| Sleep fragmentation |  | 0.204 | 0.149 | 0.999 |
| Light NREM predominance |  | 0.730 | 0.935 | 0.839 |
| N3 predominance |  | 0.630 | 0.883 | 0.888 |
| Spindle number and duration |  | 0.921 | 0.320 | 0.590 |
| REM sleep bouts |  | 0.497 | 0.155 | 0.366 |
| Respiratory disturbances |  | 0.803 | 0.241 | 0.600 |
| Slow oscillation-spindle coupling |  | 0.832 | 0.329 | 0.642 |
| Spindle amplitude |  | 0.033 | 0.962 | 0.520 |
| **Verbal Learning and Memory** | | | | |
| Sleep quantity and efficiency | 0.980 | 0.414 | 0.949 | 0.824 |
| Sleep fragmentation | 0.715 | 0.034 | 0.263 | 0.775 |
| Light NREM predominance | 0.370 | 0.560 | 0.061 | 0.585 |
| N3 predominance | 0.143 | 0.203 | 0.704 | 0.951 |
| Spindle number and duration | 0.744 | 0.224 | 0.185 | 0.875 |
| REM sleep bouts | 0.503 | 0.757 | 0.993 | 0.799 |
| Respiratory disturbances | 0.329 | 0.581 | 0.719 | 0.663 |
| Slow oscillation-spindle coupling | 0.887 | 0.599 | 0.073 | 0.610 |
| Spindle amplitude | 0.972 | 0.844 | 0.008 | 0.340 |
| **Visuospatial Function** | | | | |
| Sleep quantity and efficiency |  | 0.594 | 0.031 | 0.643 |
| Sleep fragmentation |  | 0.731 | 0.359 | 0.514 |
| Light NREM predominance |  | 0.104 | 0.278 | 0.813 |
| N3 predominance |  | 0.266 | 0.288 | 0.968 |
| Spindle number and duration |  | 0.137 | 0.229 | 0.855 |
| REM sleep bouts |  | 0.205 | 0.412 | 0.856 |
| Respiratory disturbances |  | 0.773 | 0.129 | 0.382 |
| Slow oscillation-spindle coupling |  | 0.467 | 0.458 | 0.501 |
| Spindle amplitude |  | 0.910 | 0.380 | 0.573 |

The directionality for each sleep composite was interpreted as: Sleep quantity = higher with longer sleep; Sleep fragmentation = higher with more fragmented sleep; Light NREM predominance = higher with more N1/N2 events and less REM/N3 events; N3 predominance = higher with more and longer N3; Spindle number and duration = higher value with lower number/ duration of spindles; REM sleep bouts = higher with more and shorter bouts of REM sleep; Respiratory disturbances = higher with more OSA); Spindle and slow wave coupling = higher with stronger coupling; Spindle amplitude = higher with higher amplitude.

Abbreviations: ARIC, Atherosclerosis Risk in Communities study; CHS, Cardiovascular Health Study; FHS, Framingham Heart Study; MrOS, Osteoporotic Fractures in Men Study; SOF, Study of Osteoporotic Fractures; NREM, non-rapid eye movement sleep; N3, non-rapid eye movement sleep; REM, rapid eye movement sleep.

eTable 8. Interaction p-values for sleep composite x APOE ε4 for dementia incidence

| **Sleep Composite** | **ARIC** | **CHS** | **FHS** | **MrOS** | **SOF** | **All Cohorts** |
| --- | --- | --- | --- | --- | --- | --- |
| Sleep quantity and efficiency | 0.358 | 0.604 | 0.814 | 0.352 | 0.108 | 0.898 |
| Sleep fragmentation | 0.741 | 0.413 | 0.031 | 0.124 | 0.086 | 0.333 |
| Light NREM predominance | 0.656 | 0.139 | 0.801 | 0.592 | 0.453 | 0.462 |
| N3 predominance | 0.092 | 0.028 | 0.257 | 0.816 | 0.708 | 0.397 |
| Spindle number and duration | 0.498 | 0.587 | 0.015 | 0.126 | 0.073 | 0.136 |
| REM sleep bouts | 0.427 | 0.831 | 0.501 | 0.768 | 0.518 | 0.402 |
| Respiratory disturbances | 0.232 | 0.444 | 0.944 | 0.676 | 0.435 | 0.879 |
| Slow oscillation-spindle coupling | 0.242 | 0.582 | <.001 | 0.642 | 0.716 | 0.389 |
| Spindle amplitude | 0.089 | 0.551 | 0.116 | 0.938 | 0.653 | 0.246 |

eTable 9. Interaction p-values for sleep composite x sex for dementia incidence

| **Sleep Composite** | **ARIC** | **CHS** | **FHS** | **All Cohorts** |
| --- | --- | --- | --- | --- |
| Sleep quantity and efficiency | 0.088 | 0.192 | 0.011 | 0.810 |
| Sleep fragmentation | 0.766 | 0.688 | 0.611 | 0.823 |
| Light NREM predominance | 0.950 | 0.537 | 0.149 | 0.786 |
| N3 predominance | 0.384 | 0.150 | 0.741 | 0.303 |
| Spindle number and duration | 0.079 | 0.714 | 0.281 | 0.913 |
| REM sleep bouts | 0.299 | 0.030 | 0.694 | 0.141 |
| Respiratory disturbances | 0.856 | 0.929 | 0.537 | 0.770 |
| Slow oscillation-spindle coupling | 0.987 | 0.260 | 0.356 | 0.845 |
| Spindle amplitude | 0.530 | 0.225 | 0.942 | 0.717 |

**Supplementary Methods and Figures:**

**Sleep Measures**

***Sleep macroarchitecture***

The following sleep metrics were calculated: Stage 1 (N1%), Stage 2 (N2%), Stage 3 (N3%), REM sleep (REM%), WASO (total minutes spent awake between sleep onset and offset), sleep maintenance efficiency (SME%) (total sleep time / sleep period time (the time between sleep onset and sleep offset]), total sleep time (minutes), and the apnea-hypopnea index (AHI) (defined as the number of obstructive apneas plus the number of hypopneas accompanied by a greater than 30% reduction in airflow and 4% or greater oxygen desaturation or arousal per hour of sleep). Sleep recordings in the ARIC, CHS, and FHS cohorts were limited by a maximum battery life of 9 hours, preventing examination of sleep duration greater than 9 hours across all cohorts. Thus, sleep duration was expressed as ≤6 hours vs > 6 hours (reference). As some sleep metrics were not normally distributed, transformations were applied as detailed in Table 1.

***Sleep microarchitecture***

Sleep microarchitecture was analyzed using the Luna C/C++ pipeline, developed by a member of our team (S.M.P., URL: <http://zzz.bwh.harvard.edu/luna/>). Sleep microarchitecture variables were derived from both C4/M2 and C3/M1 EEG signals. The following metrics were assessed **1) spindle metrics:** mean spindle density (no. spindles per minute), mean spindle frequency (Hz), mean spindle duration (secs), mean no. oscillations and mean spindle amplitude (μV) during N2+N3 sleep for fast (center frequency FC=15 Hz) and slow (FC=11 Hz) spindles **2) slow oscillation metrics**: slow oscillation density (no. slow oscillations per minute), mean peak-to-peak amplitude (μV), mean upward slope of the negative peak (μV/sec) and mean peak-to-peak slow oscillation duration or “wavelength” (secs) during N2+N3 sleep combined **3) slow oscillation spindle coupling metric:** phase coupling magnitude of slow and fast spindles for N2+N3 sleep.

EEG signals were resampled at 100 Hz, segmented into 30-second epochs, and then band-pass filtered with transition frequencies at 0.3 and 35Hz, using a Kaiser window zero-phase filter. For N2+N3, combined artefact free epochs were extracted of at least 10 epochs (i.e, 5 minutes) for each participant and utilized for spindle and slow oscillation detection.

All data underwent a cleaning protocol to check for outliers. The cleaning protocol consisted of two steps:

1) A 3-standard deviation (SD) rule was employed to check for outliers for each EEG signal (C4/M2 and C3/M1). If either EEG was an outlier (but not both), this recording was removed. If both observations were outliers, this data was retained at this step.
2) A similar 3-SD rule was applied for the difference between the two EEG signals. For large outlying differences, the value closest to the mean was kept and the other discarded. This way, in steps (1) and (2), no participant had their data removed completely, with all retaining at least one observation.

*Note that, where data were strictly positive (e.g., spindle count) and with a right skew,

the gamma distribution was used to identify outliers (< 0.0015 probability under the

gamma distribution, equivalent to 3-SD rule for normal distribution). Otherwise, the protocol was unchanged.

*Spindles, Slow oscillations and Slow oscillation-spindle coupling:* Spindles were detected using a modified wavelet-based approach that has been previously published in detail^33^. In brief, two classes of spindles were captured: fast (center frequency FC=15 Hz) and slow (FC=11 Hz), with spindles detected within approximately +/− 2 Hz of the target frequency. For both ‘fast’ and ‘slow’ spindles the following was calculated: mean spindle density (no. spindles per minute), mean spindle frequency, mean spindle duration, mean no. oscillations and mean spindle amplitude during N2+N3 sleep.

A heuristic was used to detect individual slow oscillations in the sleep EEG^33^. All positive-to-negative zero-crossings were identified from the filtered EEG signal. Slow oscillations were then detected using relative amplitude threshold detection. Specifically, we band-pass filtered the EEG using transition frequencies of 0.3 and 4Hz, marked all positive-to-negative zero-crossings, and designated SOs as those intervals between zero-crossings with a duration between 0.5 and 2 seconds (i.e. 0.5 to 2 Hz) and having a peak-to-peak amplitude greater than twice the average value for that individual. This algorithm was applied separately to all N2 epochs, all N3 epochs and all N2+N3 epochs combined. The following slow oscillation metrics were calculated: Slow oscillation density (no. slow oscillations per minute), mean peak-to-peak amplitude (μV), mean upward slope of the negative peak (μV/sec) and mean peak-to-peak slow oscillation duration or “wavelength” (secs) during N2+N3 sleep combined.

Slow oscillation-spindle coupling was calculated based on the Inter-Trial Phase Coherence (ITPC) statistic during combined N2+N3 sleep. The primary quantification of spindle/SO coupling was based on the phase of slow wave activity at spindle ‘peaks,’ the points of maximal spindle oscillation (peak-to-peak amplitude), typically near the spindle’s center. We estimated two measures of coupling: gross overlap and coupling magnitude. Overlap was defined as the number of spindle peaks that fell within a detected SO; we determined the null distribution of this metric empirically, by randomly shuffling spindle peaks and recalculating overlap 10,000 times. Specifically, each spindle peak was randomly shuffled only within the 30-second epoch that spanned it, to preserve any ultradian trends in spindle and/or SO rate across the night. For each individual, we normalized the overlap metric as a z-score, given the mean and standard deviation of the null distribution; we also calculated an empirical p-value for above-chance overlap.

Coupling magnitude metrics were based on the instantaneous phase from a Hilbert transform of EEG after bandpass-filtering in the 0.3 to 4 Hz range, so as not impose an artificially sinusoidal shape on SO waveforms. We calculated intra-trial phase consistency (ITPC) as a measure of the strength of spindle/SO coupling. Briefly, if the Hilbert-derived phase angle at each spindle peak is assumed to be a unit vector on a circle, with an angle matching the phase angle, then the ITPC is the magnitude of the average of these complex vectors. Under the null hypothesis of no coupling, an asymptotic p-value can be computed as exp(−n * ITPC2). Separately for fast and slow spindles, we calculated ITPC statistics for spindles with a peak that overlapped a SO. As ITPC statistics and asymptotic p-values can show bias or noise when based on a small number of spindle/SO events, or on non-sinusoidal waveforms, we generated empirical null distributions to normalize them. In order to ensure the phase coupling metric was not confounded by differences in gross spindle/SO overlap, we required null replicates to contain the same number of SO-overlapping spindle peaks, i.e. to have an identical extent of overlap. We therefore shuffled each spindle peak by a random offset, between 0 seconds and the duration of the spanning SO, wrapping as necessary. This ‘within-SO’ shuffling scheme preserved the total number of spindles, SO and their gross overlap in each null replicate, but randomized only the precise relationships between spindle peaks and SO phase.

**Sleep Bouts**

Sleep bouts were defined as a contiguous stretch of each sleep stage (N1, N2, N3 and REM). For each sleep stage, the number of bouts and the median bout duration was calculated.

**Cluster of Variables around Latent Components (CLV) analysis**

While the associations of individual sleep metrics with dementia risk are of interest, they do not exploit the correlation among sleep metrics to reveal unknown structures and incur large multiple testing penalties. To extract sub-structures from a large number of sleep metrics, and then relate them to cognitive performance and incident dementia, we first used 44 sleep metrics to identify meaningful clusters of sleep metrics using the Cluster of Variables around Latent Components (CLV) method^13^. We then can relate the latent variable representing each cluster to cognitive performance and incident dementia.

The CLV approach is related to factor analysis whereby the correlated variables are lumped together, and within each cluster, a latent (synthetic) variable is formed. A major difference between CLV and principal component analysis (PCA) or factor analysis is that a variable can only be classified into one cluster thus contributing to one latent variable in CLV, while a variable can contribute to multiple variables in PCA and factor analyses, thus the results from the latter are more difficult to interpret.

CLV employed an agglomerative technique where we started with as many clusters as the number of variables, each variable was a cluster by itself. Then in the next iteration, the number of clusters was reduced by one by aggregating two existing clusters that were chosen based on the minimum reduction of the sum of maximum eigenvalues of each cluster over all clusters before and after merging the two clusters. The iterations continued until reaching a single cluster of all variables. We first looked at the number of clusters corresponding to the first large jump in the elbow plot of merging criteria and evaluated the variance of original variables explained by all the clusters, individual cluster variances, and members of each cluster (eFigure 2). We also evaluated the performance of adjacent numbers of clusters.

eFigure 1. The pairwise correlation between age, sex and study center adjusted sleep measures

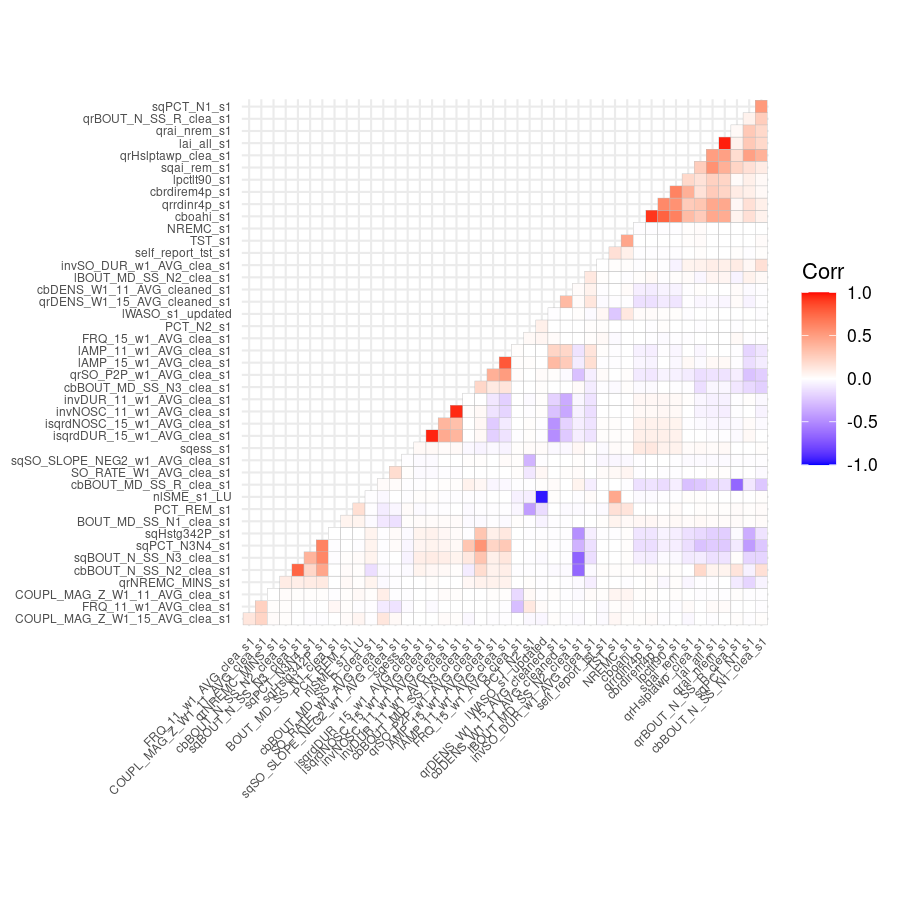

eFigure 2. Evolution of aggregation criterion with various number of clusters

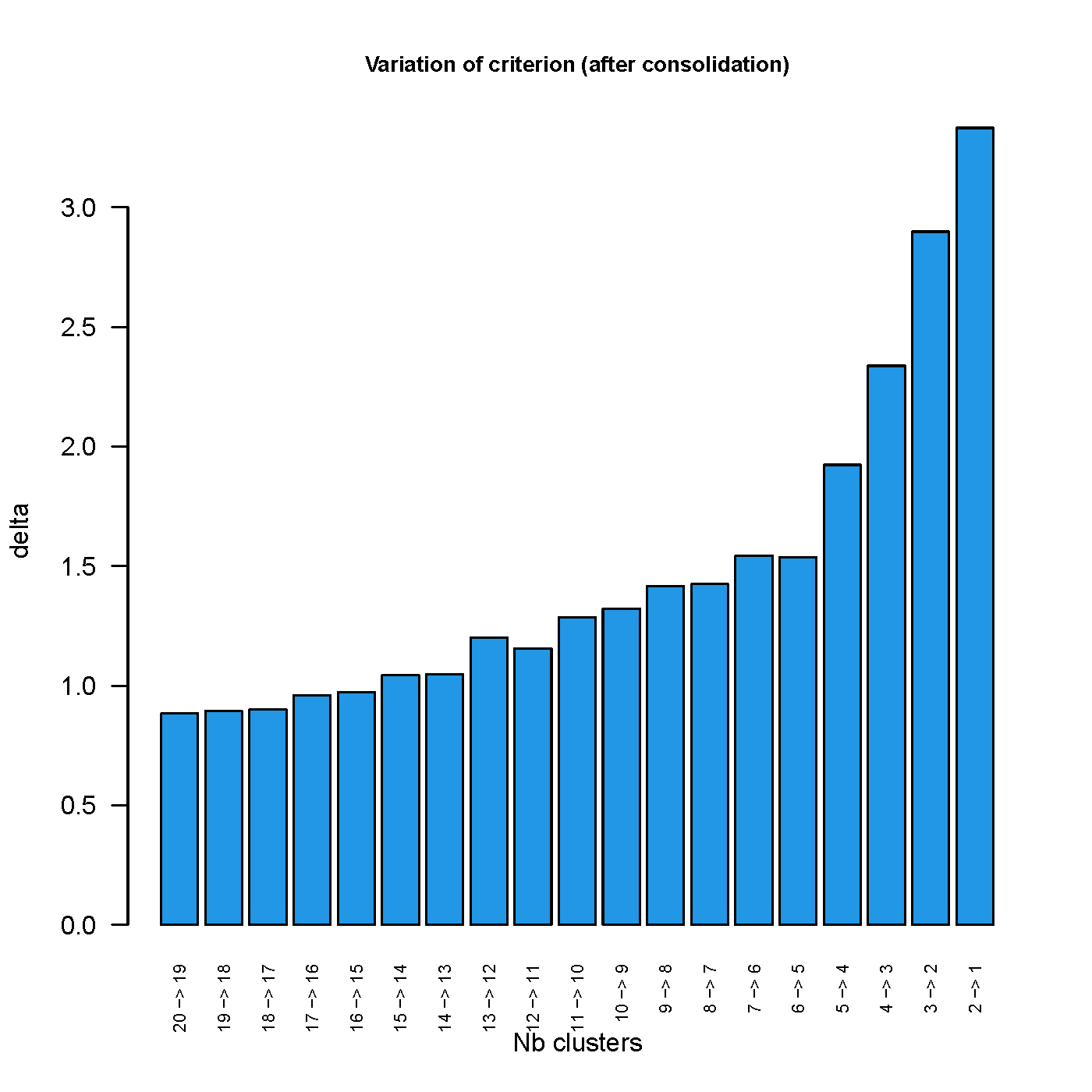

eFigure 3: Forest plots of sleep quantity and efficiency components and global cognition

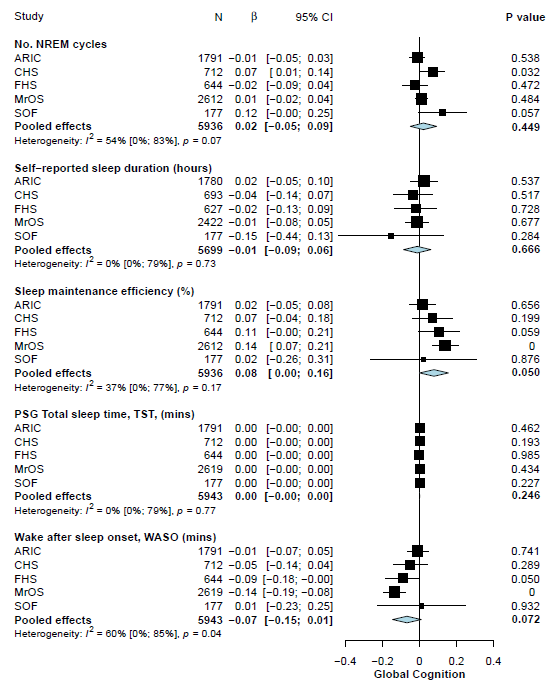

**eFigure 3:** Forest plots of the associations between sleep quantity and sleep efficiency components and global cognition. Models were adjusted for age, age^2^, sex, body mass index, anti-depressant use, sedative use, education, and the time interval between the PSG assessment and cognitive testing.

Abbreviations: ARIC, Atherosclerosis Risk in Communities study; CHS, Cardiovascular Health Study; FHS, Framingham Heart Study; MrOS, Osteoporotic Fractures in Men Study; SOF, Study of Osteoporotic Fractures.

**eFigure 4: Forest plots of slow oscillation-spindle coupling components and global cognition**

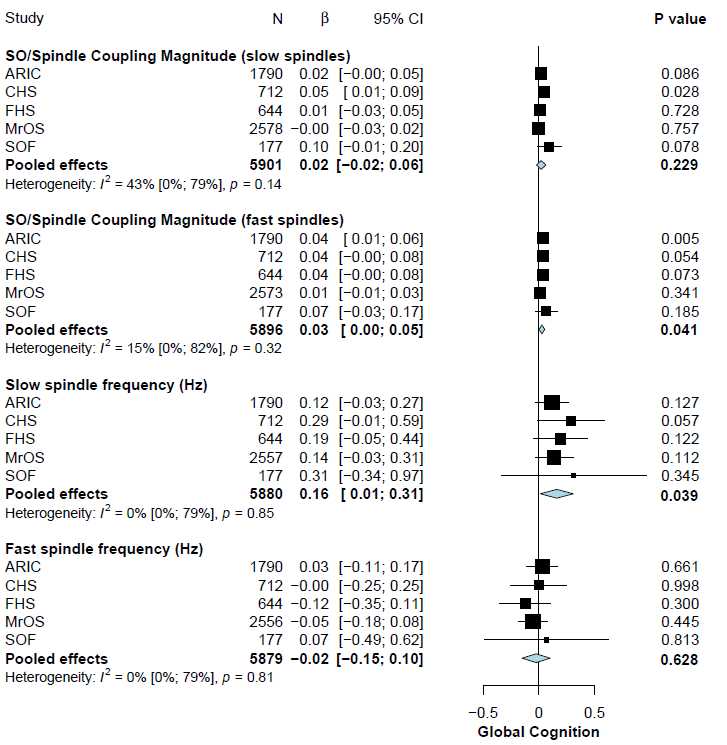

**eFigure 4:** Forest plots of the associations between sleep quantity and sleep efficiency components and global cognition. Models were adjusted for age, age^2^, sex, body mass index, anti-depressant use, sedative use, education, and the time interval between the PSG assessment and cognitive testing.

Abbreviations: ARIC, Atherosclerosis Risk in Communities study; CHS, Cardiovascular Health Study; FHS, Framingham Heart Study; MrOS, Osteoporotic Fractures in Men Study; SOF, Study of Osteoporotic Fractures.

**eFigure 5: Forest plots of sleep composites and incident dementia for model 1**
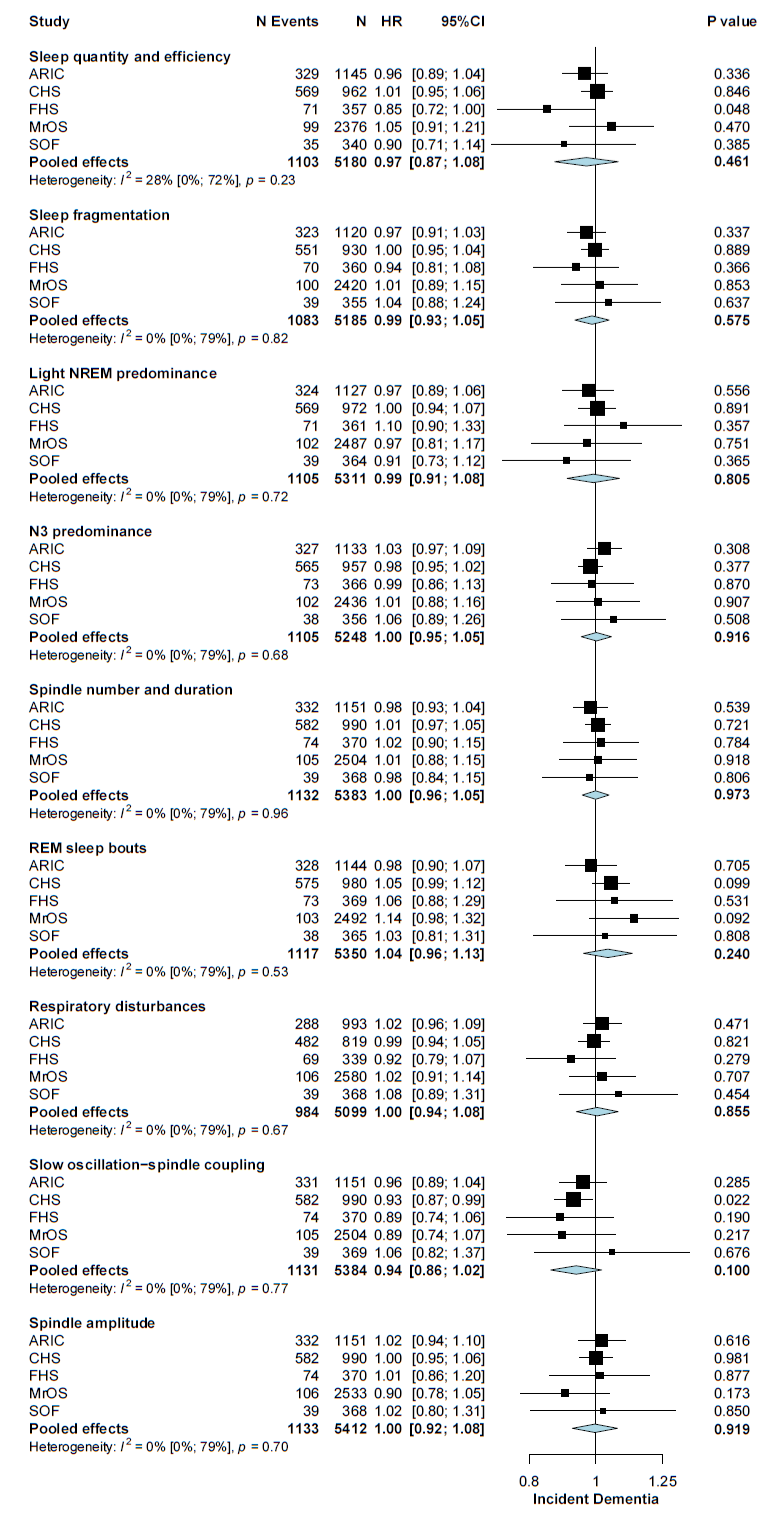

**eFigure 5:** Forest plots of the associations between sleep composites and dementia risk. Models were adjusted for age and sex. The directionality for each sleep composite was interpreted as: Sleep quantity = higher with longer sleep; Sleep fragmentation = higher with more fragmented sleep; Light NREM predominance = higher with more N1/N2 events and less REM/N3 events; N3 predominance = higher with more and longer N3; Spindle number and duration = higher value with lower number/ duration of spindles; REM sleep bouts = higher with more and shorter bouts of REM sleep; Respiratory disturbances = higher with more OSA); Spindle and slow wave coupling = higher with stronger coupling; Spindle amplitude = higher with higher amplitude.

Abbreviations: NREM, non-rapid eye movement sleep; N1, stage 1 non-rapid eye movement sleep; N2, stage 2 non-rapid eye movement sleep; N3, non-rapid eye movement sleep; REM, rapid eye movement sleep; OSA, obstructive sleep Apnea; ARIC, Atherosclerosis Risk in Communities study; CHS, Cardiovascular Health Study; FHS, Framingham Heart Study; MrOS, Osteoporotic Fractures in Men Study; SOF, Study of Osteoporotic Fractures.
